# Scoping Review: Outpatient Psychotherapeutic Care for Children and Adolescents in Germany – Status Quo and Challenges in Assessment

**DOI:** 10.1101/2024.09.12.24313544

**Authors:** Kristin Rodney-Wolf, Julian Schmitz

## Abstract

In the context of multiple global crises, including the COVID-19 pandemic, climate change, and global conflicts, children and adolescents worldwide are experiencing heightened psychological stress. As the foundation for lifelong mental health is established during childhood and adolescence, early prevention and treatment of mental health problems, such as through psychotherapy, are crucial. In Germany, current outpatient psychotherapeutic care capacities appear inadequate, while systematic evaluations of the care situation are lacking. This study investigates the state of statutory health insurance-funded outpatient psychotherapeutic care for children and adolescents in Germany and evaluates various methodological approaches for its assessment. We conducted a scoping review following the Preferred Reporting Items for Systematic Reviews and Meta-Analyses extension for Scoping Reviews (PRISMA-ScR) guidelines. Publications from January 2018 to December 2023 were sourced from PubPsych, PubMed, APA PsycInfo, Google Scholar, and ProQuest. Included studies reported quantitative primary data on the mental health of community samples of children and adolescents in Germany or their outpatient psychotherapeutic care. We included 41 publications comprising epidemiological studies, administrative data, and psychotherapist and patient reports. A lack of systematic and standardized research approaches resulted in significant variance in data. Nonetheless, qualitative analysis revealed that approximately one four children and adolescents in Germany is affected by mental health problems, while one in six to seven children and adolescents requires psychotherapeutic treatment. Yet, only up to one in fifty receives guideline-based psychotherapy. Most requests for initial psychotherapeutic consultations are unmet, with waiting times for guideline-based psychotherapy exceeding six months for at least half of the patients. Overall, our findings suggest that outpatient psychotherapeutic care for children and adolescents in Germany is still insufficient. They advocate for a systematic, multimodal, and longitudinal assessment of statutory health insurance-funded outpatient psychotherapeutic care, along with an expansion of treatment capacities to enhance access for children and adolescents in Germany.

## 1 Introduction

Worldwide, one in four to five children and adolescents suffers from a mental or behavioural disorder (1–5). With one-third to half of all mental and behavioural disorders starting in adolescence or earlier, childhood and adolescence are particularly vulnerable periods for mental health over the entire lifespan (1, 6–10). Additionally, mental and behavioural disorders are among the leading causes of disability and premature death in childhood and adolescence in industrialized nations (2, 3, 11). Currently, children and adolescents are experiencing multiple global crises and existential threats, such as the climate crisis, the aftermath of the COVID-19 pandemic, and the global effects of terror, war, and displacement. These experiences further strain the physical and mental health of children and adolescents worldwide (12–23).

Untreated mental and behavioural disorders in childhood and adolescence exhibit high rates of chronicity and persistence, along with a significant risk of developing comorbid disorders (1, 6–10). This can lead to long-term impairment in social participation and substantial secondary costs for society. Therefore, early intervention is essential to prevent mental health issues in adulthood and promote well-being and productivity throughout life. Psychotherapy is a central component of guideline-based treatment of mental and behavioural disorders in childhood and adolescence, and its effectiveness is well documented for numerous mental and behavioural disorders in these age groups (24–32). Nevertheless, even in wealthy Western countries with relatively well-developed public healthcare systems, access to professional mental health services often remains inadequate (33–35). To address this issue, the European Commission Reform Support, in cooperation with UNICEF, has recently launched a Flagship project in four EU member states to initiate reforms to expand access to mental healthcare for children and adolescents (36). To generally improve the provision of psychotherapeutic care for children and adolescents systematic and multimodal evaluations of the provision of psychotherapeutic care are needed but often lacking.

This review focusses on data regarding the outpatient psychotherapeutic care in Germany but offers a general discussion of different data sources and aims to provide an example how to evaluate access to psychotherapeutic care. Internationally, the German mental health care system for children and adolescents stands out, as it is largely funded by statutory health insurance (SHI) and offers the highest number of inpatient beds in psychotherapeutic and psychiatric units in Europe (34, 37).

However, in the outpatient sector the number of child and adolescent psychotherapists in proportion to the number of children and adolescents is lower and relatively less children and adolescents use mental health care system in Germany than in other European high-resource countries (34, 38). One reason for this is that children and adolescents with mental illnesses are often unable to find care or have to put up with unacceptably long waiting times (39, 40). Since the COVID-19 pandemic, the demand for psychotherapy, especially among children and adolescents, has increased, leading to a further rise in waiting times for psychotherapy. In 2021 and 2022, children and adolescents waited an average of around ten weeks for an initial consultation and around 25 weeks for the start of guideline-based psychotherapy (41–44). Meanwhile, SHI agencies only recorded a small increase in applications for guideline-based psychotherapy of 4–6% during the COVID-19 pandemic (45). This suggests that the supply capacities have already been fully exhausted before the pandemic and therefore, the increase in psychotherapeutic services and applications is only a poor reflection of the increased demand for psychotherapy. It is estimated that in the end only 5–10% of children and adolescents with a confirmed diagnosis of a mental or behavioural disorder receive access to guideline-based psychotherapy (32, 46, 47). “Guideline-based psychotherapy” in the German healthcare system refers to outpatient psychotherapeutic treatment funded by SHI, as regulated in the “Psychotherapy Guideline” (48). It does not include psychotherapeutic consultations and probationary sessions and can only be provided by SHI-accredited psychotherapists and psychiatrists.

Current demand planning regulations, which determine the capacity and distribution of SHI-funded outpatient healthcare in Germany, are based solely on the continuation of historical healthcare capacities rather than on empirical analyses of demand and supply (49–51). There is a lack of specific and systematic structures for psychotherapeutic care research in Germany (e.g., a longitudinal, nationwide, state-funded, and multimodal empirical monitoring), which are necessary to accurately assess the demand for mental healthcare and systematically evaluate and improve access to it (52, 53).

To accurately assess the state of psychotherapeutic care it is necessary to use valid measures for the need and demand for, the service use and provision, and the availability and accessibility of psychotherapeutic care. Therefore, different data sources are needed, such as epidemiological studies, administrative data, and reports of psychotherapist and patients themselves. Epidemiological data can be used to quantify the theoretical need for psychotherapy by estimating the prevalence of mental and behavioural disorders indicating psychotherapy. Administrative data, derived from billing data provided by SHI agencies, offer objective information on the utilisation of psychotherapeutic services over a specific period of time (e.g., profession of the service provider, coded diagnoses, billed services). Psychotherapists can furnish insights into the demand for psychotherapy (e.g., appointment requests), the need for psychotherapy (e.g., disorder diagnoses, severity of impairment, stress factors in treated patients), and their treatment capacities (e.g., waiting times). Surveys conducted among children, adolescents, or their caregivers can yield data on the self-perceived mental health, demand for psychotherapeutic treatment, and barriers to accessing psychotherapeutic care.

This review aims to provide an overview of the general epidemiological findings on mental and behavioural disorders in children and adolescents in Germany, examine the state of outpatient psychotherapeutic care for this population, and critically discuss various methodological approaches used to assess the state of child and adolescent outpatient psychotherapeutic care. Thus, the following research questions are investigated in this paper:

1. What is the current prevalence of the most common mental and behavioural disorder diagnoses and symptoms in childhood and adolescence in Germany?
2. What is the state of outpatient psychotherapy regarding need and demand, use and provision, and availability and accessibility of care according to the current database?
3. Which methods are used to assess the outpatient psychotherapeutic care for children and adolescents in Germany? Which data is particularly suitable for a valid assessment of the outpatient psychotherapeutic care situation of children and adolescents?

## 2 Methods

### 2.1 Literature Research

This review was conducted in accordance with the guidelines proposed by the Preferred Reporting Items for Systematic Reviews and Meta-Analyses extension for Scoping Reviews (PRISMA-ScR) (54). There was no registered protocol. The literature search was conducted in the PubPsych, PubMed, APA PsycInfo, and Google Scholar databases, each consulted last on 25 October 2023. Additionally, we searched in the grey literature database ProQuest on the same day, as administrative data on the mental healthcare system is often not published as empirical studies in scientific journals.

The following English search string was used to identify studies that examined the epidemiology of mental and behavioural disorders in children and adolescents or assessed the state of child and adolescent outpatient psychotherapeutic care in Germany: (((“healthcare” OR treatment) AND (mental OR psychotherap* OR psycholog*)) AND ((“health insurance” OR “claims data”) OR (patient*) OR (*therapist* OR “clinical psychologist*” OR “mental health personnel”) OR ((epidemiolog* OR prevalence OR morbidity) AND ((mental OR psycholog* OR psychopath*) AND (health OR disorder* OR status OR illness* OR depress* OR anxi* OR conduct OR internali* OR externali*))))) AND (child* OR adolescent* OR youth) AND (German* OR Baden-W*rttemberg OR Bavaria OR Berlin OR Brandenburg OR Bremen OR Hamburg OR Hesse OR Mecklenburg-Vorpommern OR “Lower Saxony” OR “North Rhine-Westphalia” OR Rhineland-Palatinate OR Saarland OR Saxony-Anhalt OR Saxony OR Schleswig-Holstein OR Thuringia).

An equivalent German search string was used: ((((Versorgung* OR Behandlung) AND (Psychotherap* OR psycholog*)) AND ((Abrechnung* OR *kasse* OR GKV OR *versicherung*) OR (Patient* OR Betroffene*) OR (*therapeut* OR “KJP” OR Versorger* OR *psycholog*))) OR ((epidemiolog* OR Pr*valenz OR H*ufigkeit*) AND (“psychische Gesundheit” OR “psychische Erkrankung*” OR “psychische St*rung*” OR psychopath* OR psycholog* OR mental* OR Depress* OR $ngst* OR Verhalten* OR internalisierend* OR externalisierend*))) AND (Kinder* OR Jugendliche*) AND ((Deutschland OR deutsch* OR “BRD” OR Bundesrepublik) OR Baden-W*rttemberg OR Bayern OR Berlin OR Brandenburg OR Bremen OR Hamburg OR Hessen OR Mecklenburg-Vorpommern OR Niedersachsen OR Nordrhein-Westfalen OR Rheinland-Pfalz OR Saarland OR Sachsen-Anhalt OR Sachsen OR Schleswig-Holstein OR Th*ringen).

For the search in Google Scholar the first 1,000 search results for the English and German search words were retrieved via “Harzing’s Publish or Perish” tool (55). English search words for Google Scholar were: outpatient, psychotherapy, care, children, adolescents, Germany. The equivalent German search terms were: ambulant, Psychotherapie, Versorgung, Kinder, Jugendliche, Deutschland. If available at the respective databases, we used automatic filters as follows: The search terms named above had to appear in the title and/or abstract of the papers and the papers had to be published between January 2018 and December 2023.

We identified additional studies via the ancestry approach by examining reference lists of studies included in this review and of reviews and meta-analyses on the same topic. The first author and a research assistant manually screened the identified studies multiple times based on titles and abstracts. Eligible studies were then textually reviewed, and the results were summarized in tables. The second author confirmed the literature search and inclusion process.

Results were filtered further using the following inclusion criteria: The publications should be written in the German or English language. They should include either epidemiological data on the mental health or data on the psychotherapeutic care of children and adolescents in Germany (i.e., administrative data, reports from psychotherapists or patients on demand, use and accessibility of psychotherapeutic care). They should further present quantitative primary data of community samples of children and adolescents.

Publications on data from other countries but Germany, data from only adult populations, clinical samples, or other specific subsamples of children and adolescents (e.g., children and adolescents of parents with mental or behavioural disorders, children and adolescents with migration history) were excluded. Further exclusion criteria comprised publications focusing solely on psychiatric, paediatric, or other medical care, healthcare cost analyses, analyses of care pathways and treatment quality, and studies reporting only on attitudes towards mental healthcare. Research evaluating specific intervention programs and methods was also excluded. Additionally, epidemiological studies on specific symptoms or syndromes not matching an ICD-10 mental or behavioural disorder diagnosis or studies in which prevalence of mental health problems was only a secondary outcome met exclusion criteria. Opinion papers, statements, comments, reviews, and meta-analyses without original primary data were not included. If data was both analysed on the state and federal level, state reports were excluded if their data was included in a federal report (e.g. “DAK Gesundheitsreport”). If multiple publications reported on the same data (e.g., studies published in English and in German), they were regarded as one publication but all versions were cited. The process of study selection is depicted in Figure 1.

**Figure 1.**
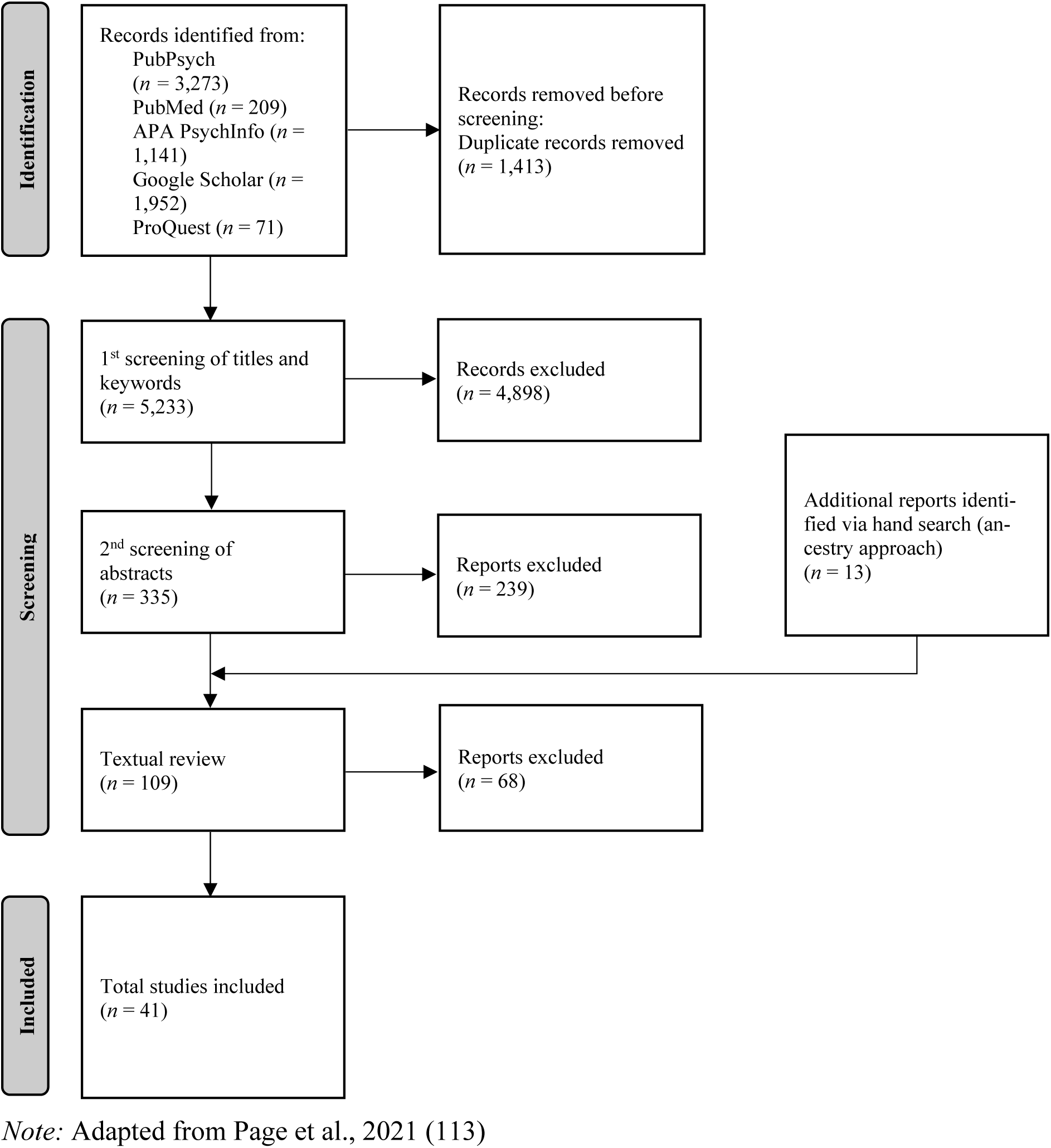
Process and Results of Literature Research

### 2.2 Synthesis of Results

In order to critically examine the suitability and investigated constructs of different methods to assess the state of the outpatient psychotherapeutic care for children and adolescents, the included studies were sorted into four categories according to data source or research focus: epidemiological studies, administrative studies, psychotherapists’ reports, and patients’ reports. The latter could include assessments of the psychotherapeutic care by the potential patients’ themselves (i.e. children and adolescents) or their caregivers. If a study reported on both mental health problems and psychotherapeutic care from the point of view of children, adolescents, and/or caregivers, it was listed in both categories, epidemiological data and patient reports. Epidemiological studies were specifically sought to answer the first research question. All data sources were assessed to answer the second and third research question. If applicable, data on epidemiology of mental and behavioural disorders and on the provision of psychotherapeutic care was summarized separately for each data source. We sorted epidemiological studies by the age of the assessed children and adolescents (< six years, six–12 years, > 12 years) to account for age effects in the prevalence and incidence of mental and behavioural disorder diagnoses and symptoms. As the COVID-19 pandemic led to both a disruption of the healthcare system and an increase of mental distress in children and adolescents, data on the epidemiology of mental and behavioural disorder diagnoses and symptoms and on the provision of mental healthcare from before (i.e., until 2020) and during the COVID-19 pandemic (i.e., from 2020) were analysed separately.

## 3 Results

### 3.1 Overview of Included Studies

In total, we included 41 studies, published between April 2018 and November 2023. The included studies were summarized in four tables: Table 1 displays the 18 epidemiological studies. Table 2 depicts the 15 publications on administrative data. Table 3 includes the seven studies on psychotherapists’ reports and Table 4 summarizes the remaining four studies on patient reports. Three studies were listed in two categories (56–58) because they contained information on both epidemiology and mental healthcare provision from the patients’ perspective.

**Table 1.**
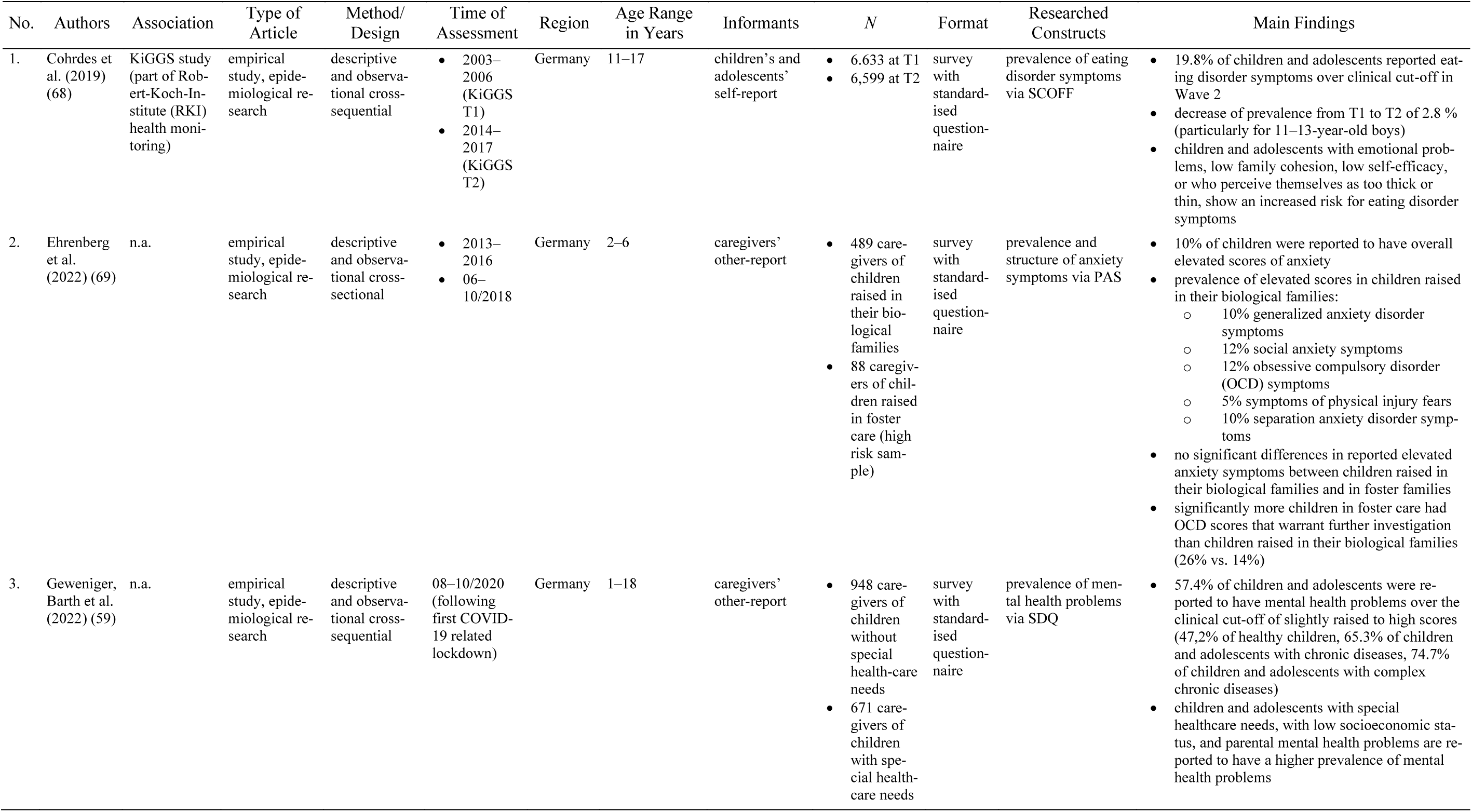
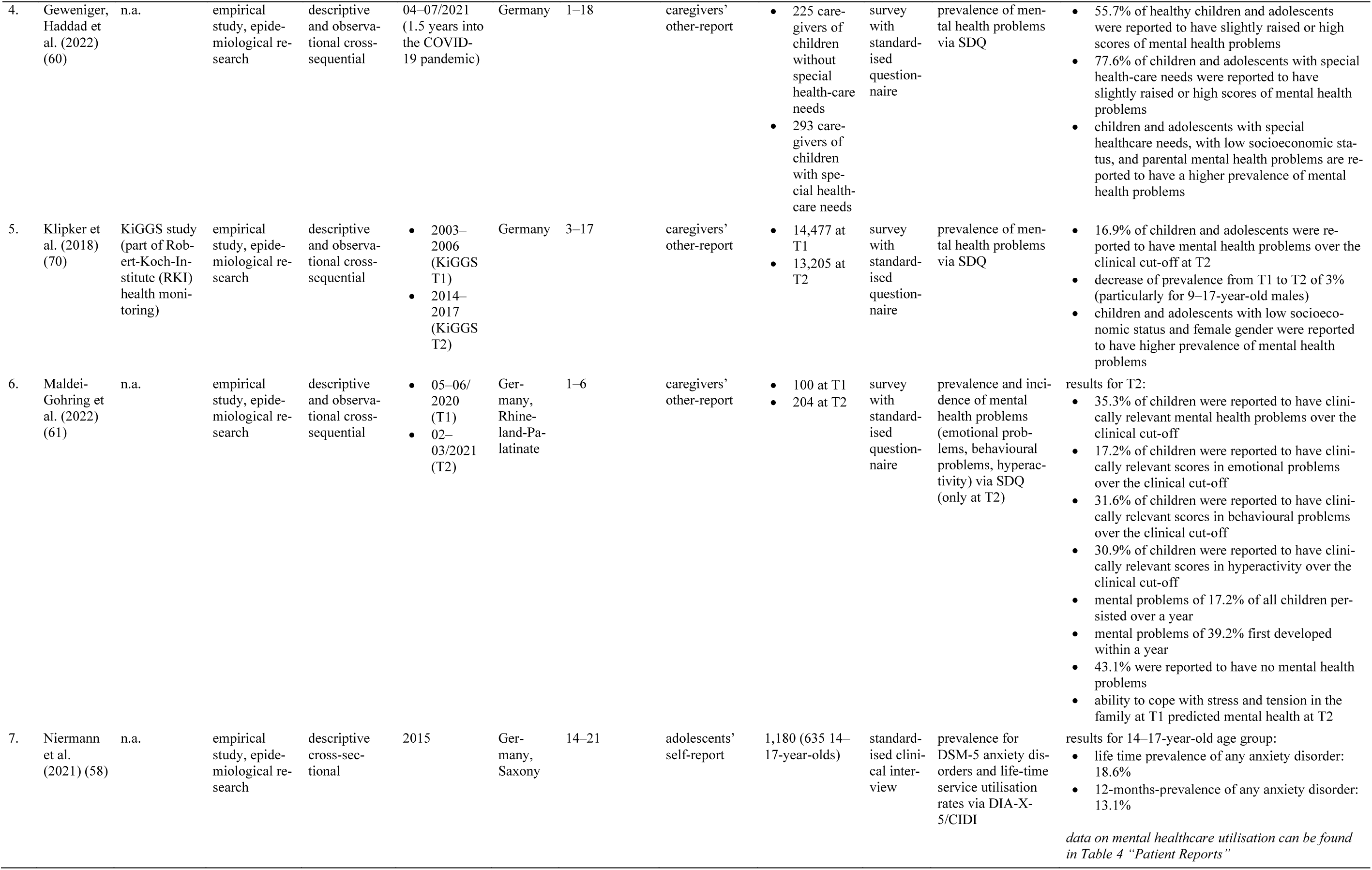
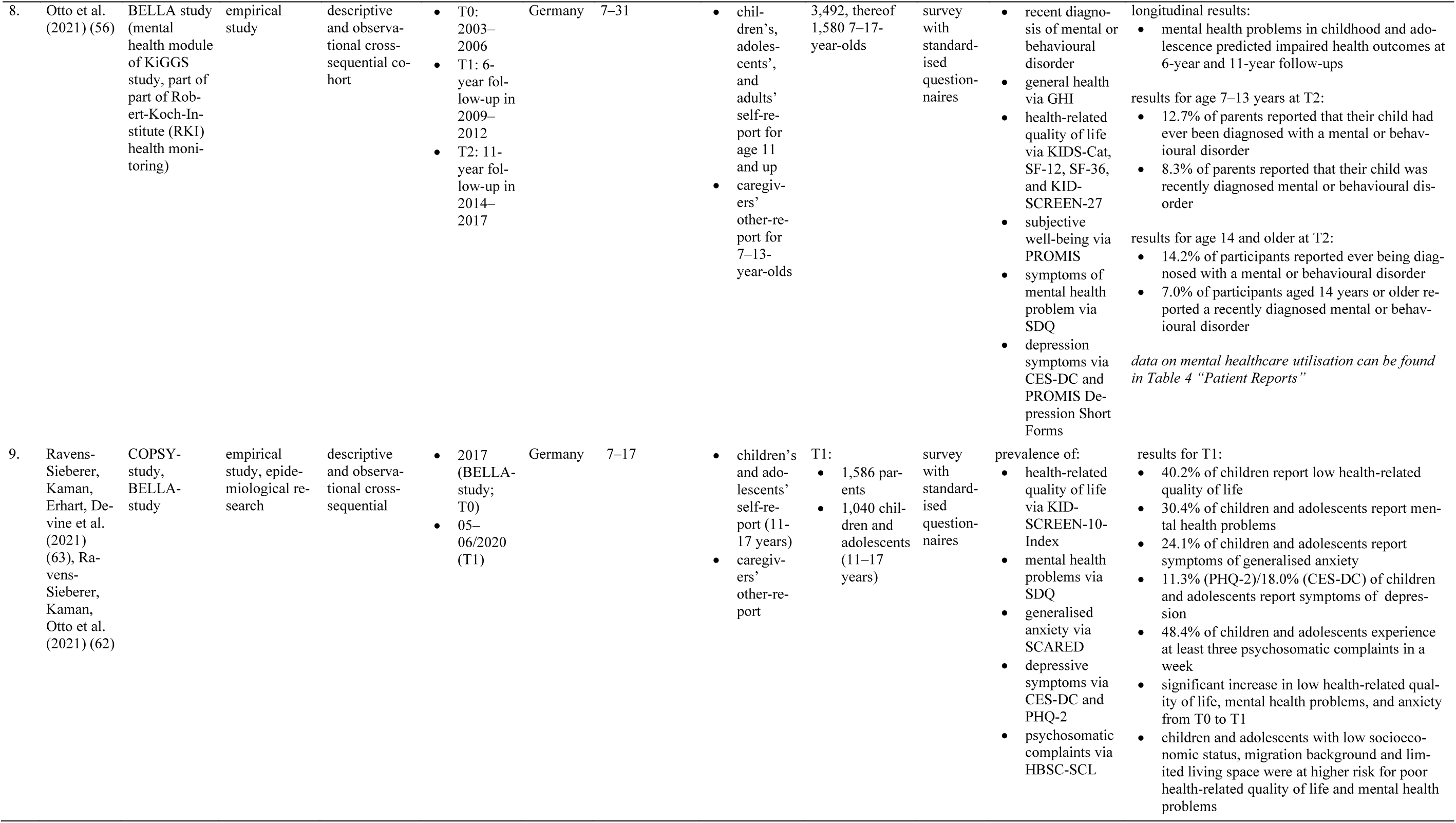
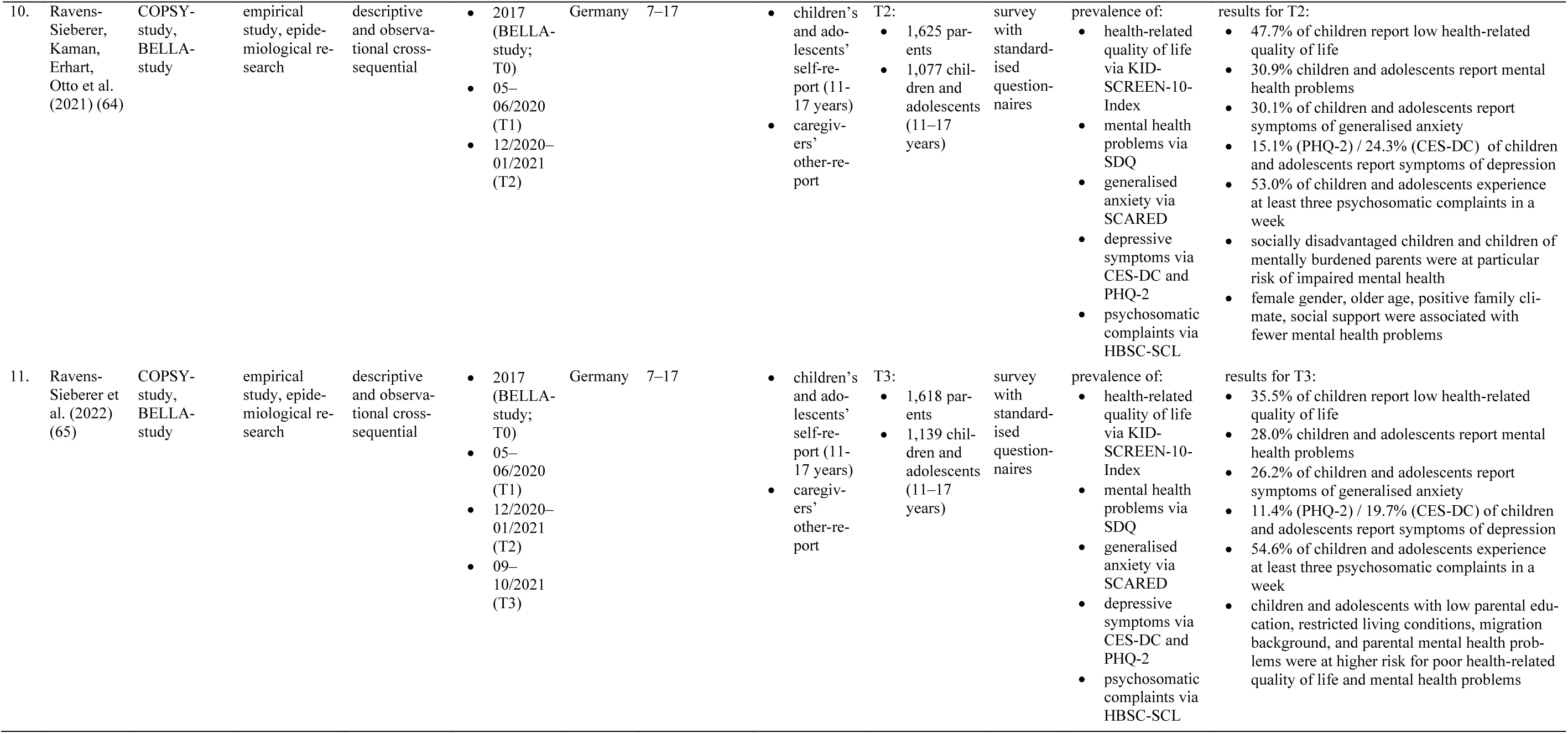
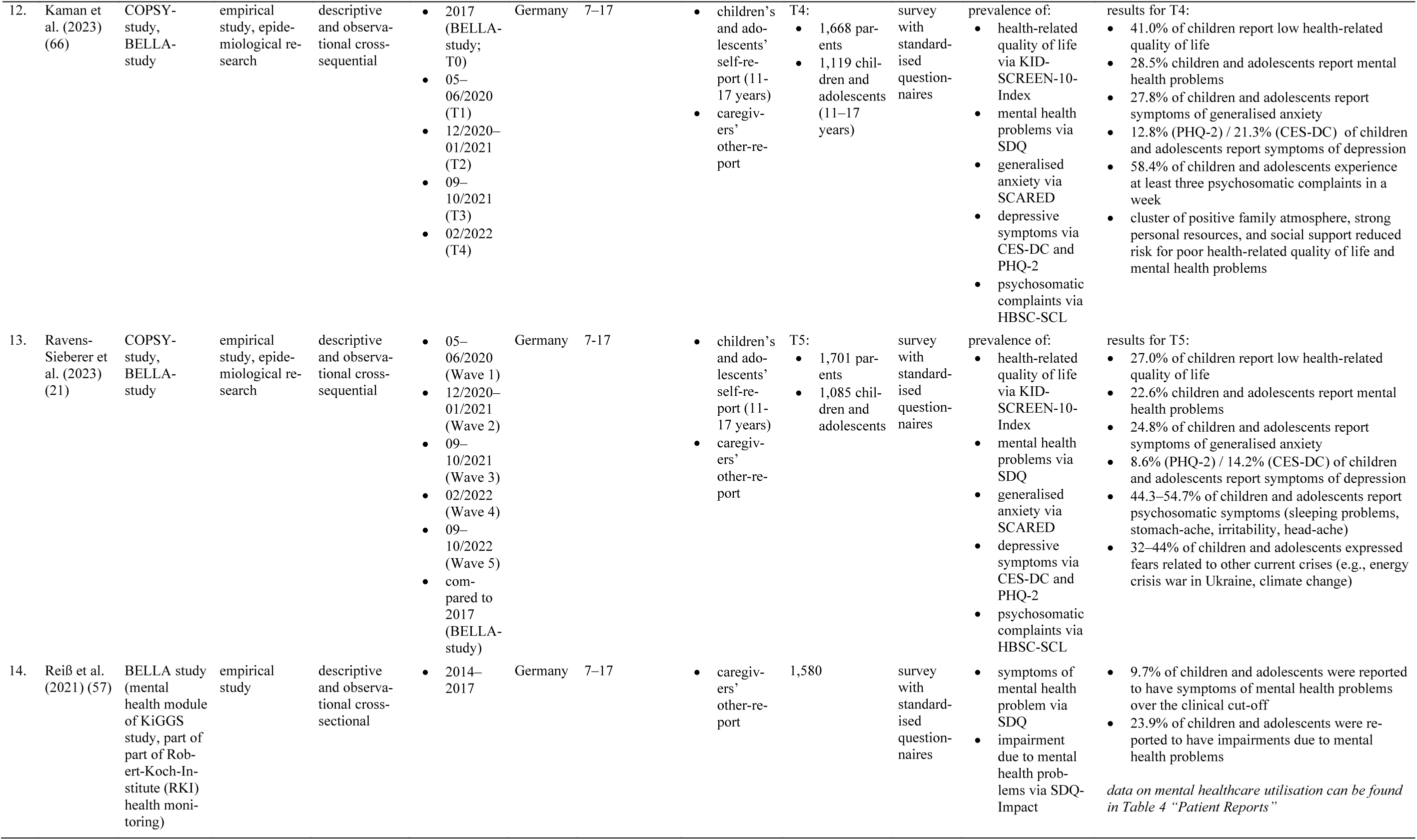
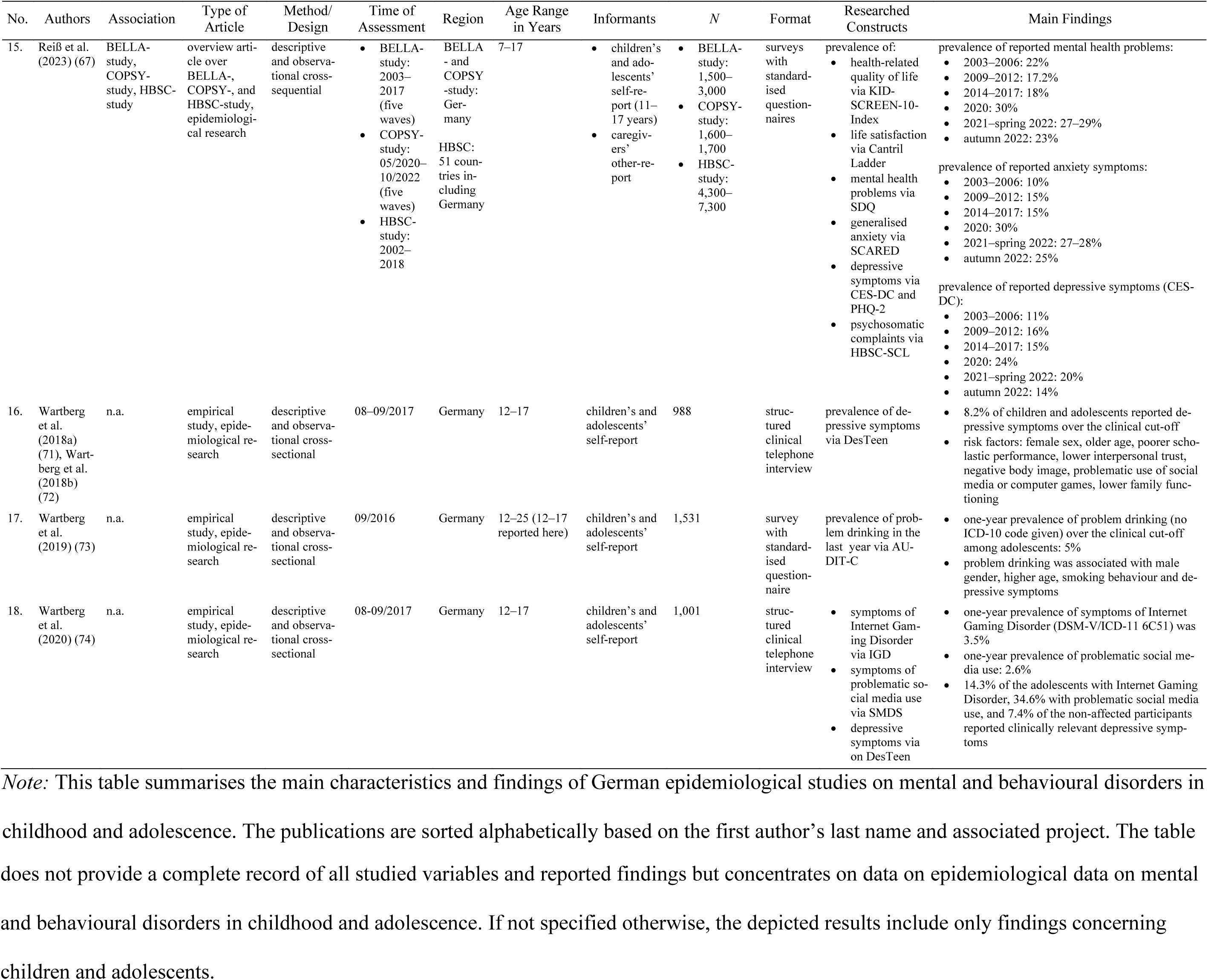
Overview of Epidemiological Studies.

| No. | Authors | Association | Type of Article | Method/Design | Time of Assessment | Region | Age Range in Years | Informants | N | Format | Researched Constructs | Main Findings |
| --- | --- | --- | --- | --- | --- | --- | --- | --- | --- | --- | --- | --- |
| 1. | Cohrdes et al. (2019) (68) | KiGGS study (part of Robert-Koch-Institute (RKI) health monitoring) | empirical study, epidemiological research | descriptive and observational cross-sequential | • 2003–2006 (KiGGS T1)<br>• 2014–2017 (KiGGS T2) | Germany | 11–17 | children’s and adolescents’ self-report | • 6,633 at T1<br>• 6,599 at T2 | survey with standardised questionnaire | prevalence of eating disorder symptoms via SCOFF | • 19.8% of children and adolescents reported eating disorder symptoms over clinical cut-off in Wave 2<br>• decrease of prevalence from T1 to T2 of 2.8 % (particularly for 11–13-year-old boys)<br>• children and adolescents with emotional problems, low family cohesion, low self-efficacy, or who perceive themselves as too thick or thin, show an increased risk for eating disorder symptoms |
| 2. | Ehrenberg et al. (2022) (69) | n.a. | empirical study, epidemiological research | descriptive and observational cross-sectional | • 2013–2016<br>• 06–10/2018 | Germany | 2–6 | caregivers’ other-report | • 489 caregivers of children raised in their biological families<br>• 88 caregivers of children raised in foster care (high risk sample) | survey with standardised questionnaire | prevalence and structure of anxiety symptoms via PAS | • 10% of children were reported to have overall elevated scores of anxiety<br>• prevalence of elevated scores in children raised in their biological families:<br>◦ 10% generalized anxiety disorder symptoms<br>◦ 12% social anxiety symptoms<br>◦ 12% obsessive compulsory disorder (OCD) symptoms<br>◦ 5% symptoms of physical injury fears<br>◦ 10% separation anxiety disorder symptoms<br>• no significant differences in reported elevated anxiety symptoms between children raised in their biological families and in foster families<br>• significantly more children in foster care had OCD scores that warrant further investigation than children raised in their biological families (26% vs. 14%) |
| 3. | Geweniger, Barth et al. (2022) (59) | n.a. | empirical study, epidemiological research | descriptive and observational cross-sequential | 08–10/2020 (following first COVID-19 related lockdown) | Germany | 1–18 | caregivers’ other-report | • 948 caregivers of children without special health-care needs<br>• 671 caregivers of children with special health-care needs | survey with standardised questionnaire | prevalence of mental health problems via SDQ | • 57.4% of children and adolescents were reported to have mental health problems over the clinical cut-off of slightly raised to high scores (47.2% of healthy children, 65.3% of children and adolescents with chronic diseases, 74.7% of children and adolescents with complex chronic diseases)<br>• children and adolescents with special healthcare needs, with low socioeconomic status, and parental mental health problems are reported to have a higher prevalence of mental health problems |
| 4. | Geweniger, Haddad et al. (2022) (60) | n.a. | empirical study, epidemiological research | descriptive and observational cross-sequential | 04–07/2021 (1.5 years into the COVID-19 pandemic) | Germany | 1–18 | caregivers' other-report | 225 caregivers of children without special health-care needs<br>293 caregivers of children with special health-care needs | survey with standardised questionnaire | prevalence of mental health problems via SDQ | 55.7% of healthy children and adolescents were reported to have slightly raised or high scores of mental health problems<br>77.6% of children and adolescents with special health-care needs were reported to have slightly raised or high scores of mental health problems<br>children and adolescents with special healthcare needs, with low socioeconomic status, and parental mental health problems are reported to have a higher prevalence of mental health problems |
| 5. | Klipker et al. (2018) (70) | KiGGS study (part of Robert-Koch-Institute (RKI) health monitoring) | empirical study, epidemiological research | descriptive and observational cross-sequential | 2003–2006 (KiGGS T1)<br>2014–2017 (KiGGS T2) | Germany | 3–17 | caregivers' other-report | 14,477 at T1<br>13,205 at T2 | survey with standardised questionnaire | prevalence of mental health problems via SDQ | 16.9% of children and adolescents were reported to have mental health problems over the clinical cut-off at T2<br>decrease of prevalence from T1 to T2 of 3% (particularly for 9–17-year-old males)<br>children and adolescents with low socioeconomic status and female gender were reported to have higher prevalence of mental health problems |
| 6. | Maldei-Gohring et al. (2022) (61) | n.a. | empirical study, epidemiological research | descriptive and observational cross-sequential | 05–06/2020 (T1)<br>02–03/2021 (T2) | Germany, Rhineland-Palatinate | 1–6 | caregivers' other-report | 100 at T1<br>204 at T2 | survey with standardised questionnaire | prevalence and incidence of mental health problems (emotional problems, behavioural problems, hyperactivity) via SDQ (only at T2) | results for T2:<br>35.3% of children were reported to have clinically relevant mental health problems over the clinical cut-off<br>17.2% of children were reported to have clinically relevant scores in emotional problems over the clinical cut-off<br>31.6% of children were reported to have clinically relevant scores in behavioural problems over the clinical cut-off<br>30.9% of children were reported to have clinically relevant scores in hyperactivity over the clinical cut-off<br>mental problems of 17.2% of all children persisted over a year<br>mental problems of 39.2% first developed within a year<br>43.1% were reported to have no mental health problems<br>ability to cope with stress and tension in the family at T1 predicted mental health at T2 |
| 7. | Niermann et al. (2021) (58) | n.a. | empirical study, epidemiological research | descriptive cross-sectional | 2015 | Germany, Saxony | 14–21 | adolescents' self-report | 1,180 (635 14–17-year-olds) | standardised clinical interview | prevalence for DSM-5 anxiety disorders and life-time service utilisation rates via DIA-X-5/CIDI | results for 14–17-year-old age group:<br>life time prevalence of any anxiety disorder: 18.6%<br>12-months-prevalence of any anxiety disorder: 13.1%<br>*data on mental healthcare utilisation can be found in Table 4 “Patient Reports”* |
| 8. | Otto et al. (2021) (56) | BELLA study (mental health module of KiGGS study, part of part of Robert-Koch-Institute (RKI) health monitoring) | empirical study | descriptive and observational cross-sequential cohort | T0: 2003–2006<br>T1: 6-year follow-up in 2009–2012<br>T2: 11-year follow-up in 2014–2017 | Germany | 7–31 | children's, adolescents', and adults' self-report for age 11 and up<br>caregivers' other-report for 7–13-year-olds | 3,492, thereof 1,580 7–17-year-olds | survey with standardised questionnaires | recent diagnosis of mental or behavioural disorder<br>general health via GHI<br>health-related quality of life via KIDS-Cat, SF-12, SF-36, and KID-SCREEN-27<br>subjective well-being via PROMIS<br>symptoms of mental health problem via SDQ<br>depression symptoms via CES-DC and PROMIS Depression Short Forms | longitudinal results:<br>mental health problems in childhood and adolescence predicted impaired health outcomes at 6-year and 11-year follow-ups<br>results for age 7–13 years at T2:<br>12.7% of parents reported that their child had ever been diagnosed with a mental or behavioural disorder<br>8.3% of parents reported that their child was recently diagnosed mental or behavioural disorder<br>results for age 14 and older at T2:<br>14.2% of participants reported ever being diagnosed with a mental or behavioural disorder<br>7.0% of participants aged 14 years or older reported a recently diagnosed mental or behavioural disorder<br>*data on mental healthcare utilisation can be found in Table 4 "Patient Reports"* |
| 9. | Ravens-Sieberer, Kaman, Erhart, Devine et al. (2021) (63), Ravens-Sieberer, Kaman, Otto et al. (2021) (62) | COPSY-study, BELLA-study | empirical study, epidemiological research | descriptive and observational cross-sequential | 2017 (BELLA-study; T0)<br>05–06/2020 (T1) | Germany | 7–17 | children's and adolescents' self-report (11–17 years)<br>caregivers' other-report | T1:<br>1,586 parents<br>1,040 children and adolescents (11–17 years) | survey with standardised questionnaires | prevalence of:<br>health-related quality of life via KID-SCREEN-10-Index<br>mental health problems via SDQ<br>generalised anxiety via SCARED<br>depressive symptoms via CES-DC and PHQ-2<br>psychosomatic complaints via HBSC-SCL | results for T1:<br>40.2% of children report low health-related quality of life<br>30.4% of children and adolescents report mental health problems<br>24.1% of children and adolescents report symptoms of generalised anxiety<br>11.3% (PHQ-2)/18.0% (CES-DC) of children and adolescents report symptoms of depression<br>48.4% of children and adolescents experience at least three psychosomatic complaints in a week<br>significant increase in low health-related quality of life, mental health problems, and anxiety from T0 to T1<br>children and adolescents with low socioeconomic status, migration background and limited living space were at higher risk for poor health-related quality of life and mental health problems |
| 10. | Ravens-Sieberer, Kaman, Erhart, Otto et al. (2021) (64) | COPSY-study, BELLA-study | empirical study, epidemiological research | descriptive and observational cross-sequential | 2017 (BELLA-study; T0)<br>05–06/2020 (T1)<br>12/2020–01/2021 (T2) | Germany | 7–17 | children's and adolescents' self-report (11–17 years)<br>caregivers' other-report | T2:<br>1,625 parents<br>1,077 children and adolescents (11–17 years) | survey with standardised questionnaires | prevalence of:<br>health-related quality of life via KID-SCREEN-10-Index<br>mental health problems via SDQ<br>generalised anxiety via SCARED<br>depressive symptoms via CES-DC and PHQ-2<br>psychosomatic complaints via HBSC-SCL | results for T2:<br>47.7% of children report low health-related quality of life<br>30.9% children and adolescents report mental health problems<br>30.1% of children and adolescents report symptoms of generalised anxiety<br>15.1% (PHQ-2) / 24.3% (CES-DC) of children and adolescents report symptoms of depression<br>53.0% of children and adolescents experience at least three psychosomatic complaints in a week<br>socially disadvantaged children and children of mentally burdened parents were at particular risk of impaired mental health<br>female gender, older age, positive family climate, social support were associated with fewer mental health problems |
| 11. | Ravens-Sieberer et al. (2022) (65) | COPSY-study, BELLA-study | empirical study, epidemiological research | descriptive and observational cross-sequential | 2017 (BELLA-study; T0)<br>05–06/2020 (T1)<br>12/2020–01/2021 (T2)<br>09–10/2021 (T3) | Germany | 7–17 | children's and adolescents' self-report (11–17 years)<br>caregivers' other-report | T3:<br>1,618 parents<br>1,139 children and adolescents (11–17 years) | survey with standardised questionnaires | prevalence of:<br>health-related quality of life via KID-SCREEN-10-Index<br>mental health problems via SDQ<br>generalised anxiety via SCARED<br>depressive symptoms via CES-DC and PHQ-2<br>psychosomatic complaints via HBSC-SCL | results for T3:<br>35.5% of children report low health-related quality of life<br>28.0% children and adolescents report mental health problems<br>26.2% of children and adolescents report symptoms of generalised anxiety<br>11.4% (PHQ-2) / 19.7% (CES-DC) of children and adolescents report symptoms of depression<br>54.6% of children and adolescents experience at least three psychosomatic complaints in a week<br>children and adolescents with low parental education, restricted living conditions, migration background, and parental mental health problems were at higher risk for poor health-related quality of life and mental health problems |

| No. | Authors | Association | Type of Article | Method/ Design | Time of Assessment | Region | Age Range in Years | Informants | N | Format | Researched Constructs | Main Findings |
| --- | --- | --- | --- | --- | --- | --- | --- | --- | --- | --- | --- | --- |
| 12. | Kaman et al. (2023) (66) | COPSY-study, BELLA-study | empirical study, epidemiological research | descriptive and observational cross-sequential | 2017 (BELLA-study; T0)<br>05–06/2020 (T1)<br>12/2020–01/2021 (T2)<br>09–10/2021 (T3)<br>02/2022 (T4) | Germany | 7–17 | children's and adolescents' self-report (11–17 years)<br>caregivers' other-report | T4:<br>1,668 parents<br>1,119 children and adolescents (11–17 years) | survey with standardised questionnaires | prevalence of:<br>health-related quality of life via KID-SCREEN-10-Index<br>mental health problems via SDQ<br>generalised anxiety via SCARED<br>depressive symptoms via CES-DC and PHQ-2<br>psychosomatic complaints via HBSC-SCL | results for T4:<br>41.0% of children report low health-related quality of life<br>28.5% children and adolescents report mental health problems<br>27.8% of children and adolescents report symptoms of generalised anxiety<br>12.8% (PHQ-2) / 21.3% (CES-DC) of children and adolescents report symptoms of depression<br>58.4% of children and adolescents experience at least three psychosomatic complaints in a week<br>cluster of positive family atmosphere, strong personal resources, and social support reduced risk for poor health-related quality of life and mental health problems |
| 13. | Ravens-Sieberer et al. (2023) (21) | COPSY-study, BELLA-study | empirical study, epidemiological research | descriptive and observational cross-sequential | 05–06/2020 (Wave 1)<br>12/2020–01/2021 (Wave 2)<br>09–10/2021 (Wave 3)<br>02/2022 (Wave 4)<br>09–10/2022 (Wave 5)<br>compared to 2017 (BELLA-study) | Germany | 7–17 | children's and adolescents' self-report (11–17 years)<br>caregivers' other-report | T5:<br>1,701 parents<br>1,085 children and adolescents | survey with standardised questionnaires | prevalence of:<br>health-related quality of life via KID-SCREEN-10-Index<br>mental health problems via SDQ<br>generalised anxiety via SCARED<br>depressive symptoms via CES-DC and PHQ-2<br>psychosomatic complaints via HBSC-SCL | results for T5:<br>27.0% of children report low health-related quality of life<br>22.6% children and adolescents report mental health problems<br>24.8% of children and adolescents report symptoms of generalised anxiety<br>8.6% (PHQ-2) / 14.2% (CES-DC) of children and adolescents report symptoms of depression<br>44.3–54.7% of children and adolescents report psychosomatic symptoms (sleeping problems, stomach-ache, irritability, head-ache)<br>32–44% of children and adolescents expressed fears related to other current crises (e.g., energy crisis war in Ukraine, climate change) |
| 14. | Reiß et al. (2021) (57) | BELLA study (mental health module of KiGGS study, part of part of Robert-Koch-Institute (RKI) health monitoring) | empirical study | descriptive and observational cross-sectional | 2014–2017 | Germany | 7–17 | caregivers' other-report | 1,580 | survey with standardised questionnaires | symptoms of mental health problem via SDQ<br>impairment due to mental health problems via SDQ-Impact | 9.7% of children and adolescents were reported to have symptoms of mental health problems over the clinical cut-off<br>23.9% of children and adolescents were reported to have impairments due to mental health problems<br>*data on mental healthcare utilisation can be found in Table 4 “Patient Reports”* |
| 15. | Reiß et al. (2023) (67) | BELLA-study, COPSYS-study, HBSC-study | overview article over BELLA-, COPSYS-, and HBSC-study, epidemiological research | descriptive and observational cross-sequential | BELLA-study: 2003–2017 (five waves)<br>COPSYS-study: 05/2020–10/2022 (five waves)<br>HBSC-study: 2002–2018 | BELLA- and COPSYS-study: Germany<br>HBSC: 51 countries including Germany | 7–17 | children's and adolescents' self-report (11–17 years)<br>caregivers' other-report | BELLA-study: 1,500–3,000<br>COPSYS-study: 1,600–1,700<br>HBSC-study: 4,300–7,300 | surveys with standardised questionnaires | prevalence of:<br>health-related quality of life via KID-SCREEN-10-Index<br>life satisfaction via Cantril Ladder<br>mental health problems via SDQ<br>generalised anxiety via SCARED<br>depressive symptoms via CES-DC and PHQ-2<br>psychosomatic complaints via HBSC-SCL | prevalence of reported mental health problems:<br>2003–2006: 22%<br>2009–2012: 17.2%<br>2014–2017: 18%<br>2020: 30%<br>2021–spring 2022: 27–29%<br>autumn 2022: 23%<br>prevalence of reported anxiety symptoms:<br>2003–2006: 10%<br>2009–2012: 15%<br>2014–2017: 15%<br>2020: 30%<br>2021–spring 2022: 27–28%<br>autumn 2022: 25%<br>prevalence of reported depressive symptoms (CES-DC):<br>2003–2006: 11%<br>2009–2012: 16%<br>2014–2017: 15%<br>2020: 24%<br>2021–spring 2022: 20%<br>autumn 2022: 14% |
| 16. | Wartberg et al. (2018a) (71), Wartberg et al. (2018b) (72) | n.a. | empirical study, epidemiological research | descriptive and observational cross-sectional | 08–09/2017 | Germany | 12–17 | children's and adolescents' self-report | 988 | structured clinical telephone interview | prevalence of depressive symptoms via DesTeen | 8.2% of children and adolescents reported depressive symptoms over the clinical cut-off<br>risk factors: female sex, older age, poorer scholastic performance, lower interpersonal trust, negative body image, problematic use of social media or computer games, lower family functioning |
| 17. | Wartberg et al. (2019) (73) | n.a. | empirical study, epidemiological research | descriptive and observational cross-sectional | 09/2016 | Germany | 12–25 (12–17 reported here) | children's and adolescents' self-report | 1,531 | survey with standardised questionnaire | prevalence of problem drinking in the last year via AUDIT-C | one-year prevalence of problem drinking (no ICD-10 code given) over the clinical cut-off among adolescents: 5%<br>problem drinking was associated with male gender, higher age, smoking behaviour and depressive symptoms |
| 18. | Wartberg et al. (2020) (74) | n.a. | empirical study, epidemiological research | descriptive and observational cross-sectional | 08-09/2017 | Germany | 12–17 | children's and adolescents' self-report | 1,001 | structured clinical telephone interview | symptoms of Internet Gaming Disorder via IGD<br>symptoms of problematic social media use via SMDS<br>depressive symptoms via on DesTeen | one-year prevalence of symptoms of Internet Gaming Disorder (DSM-V/ICD-11 6C51) was 3.5%<br>one-year prevalence of problematic social media use: 2.6%<br>14.3% of the adolescents with Internet Gaming Disorder, 34.6% with problematic social media use, and 7.4% of the non-affected participants reported clinically relevant depressive symptoms |

*Note:* This table summarises the main characteristics and findings of German epidemiological studies on mental and behavioural disorders in childhood and adolescence. The publications are sorted alphabetically based on the first author's last name and associated project. The table
does not provide a complete record of all studied variables and reported findings but concentrates on data on epidemiological data on mental and behavioural disorders in childhood and adolescence. If not specified otherwise, the depicted results include only findings concerning children and adolescents.

**Table 2.**
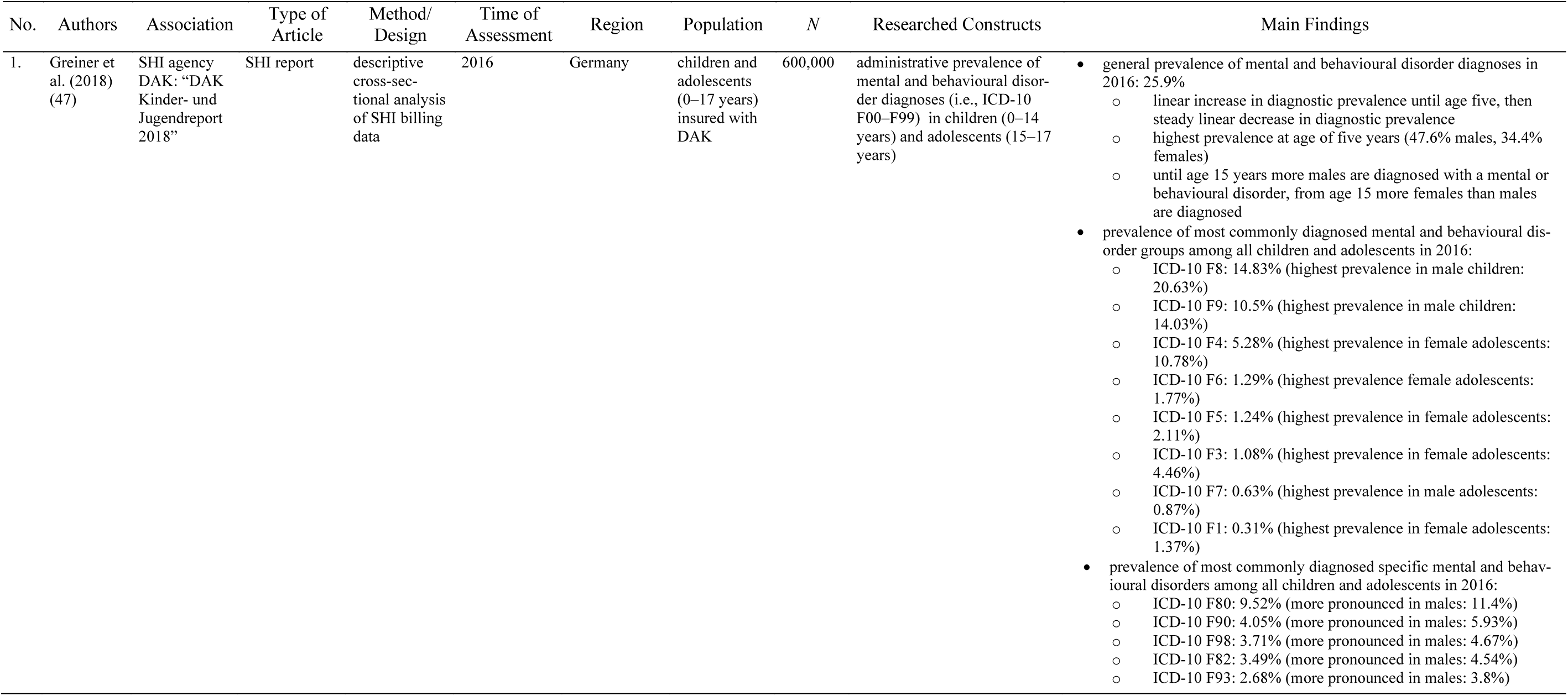
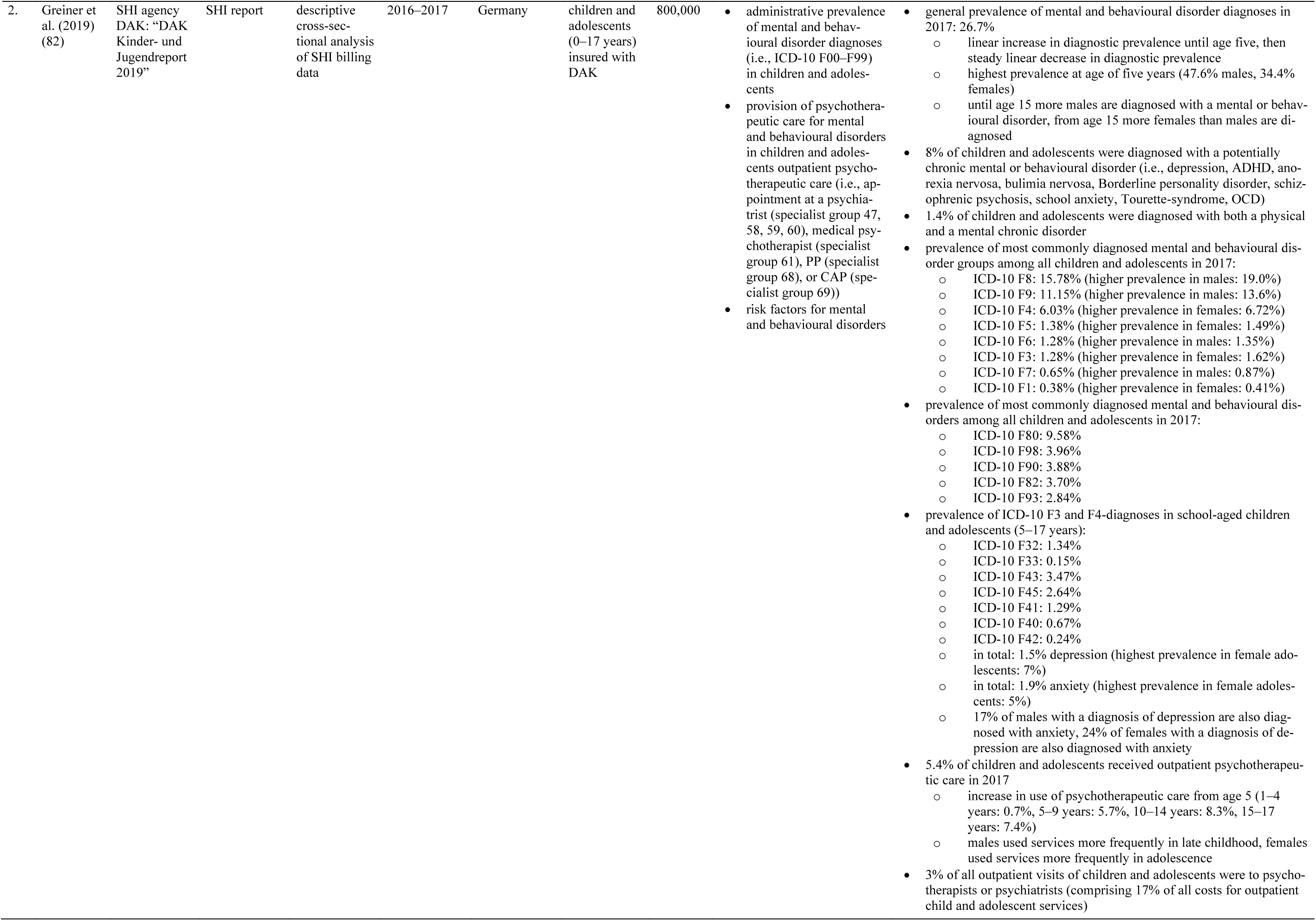
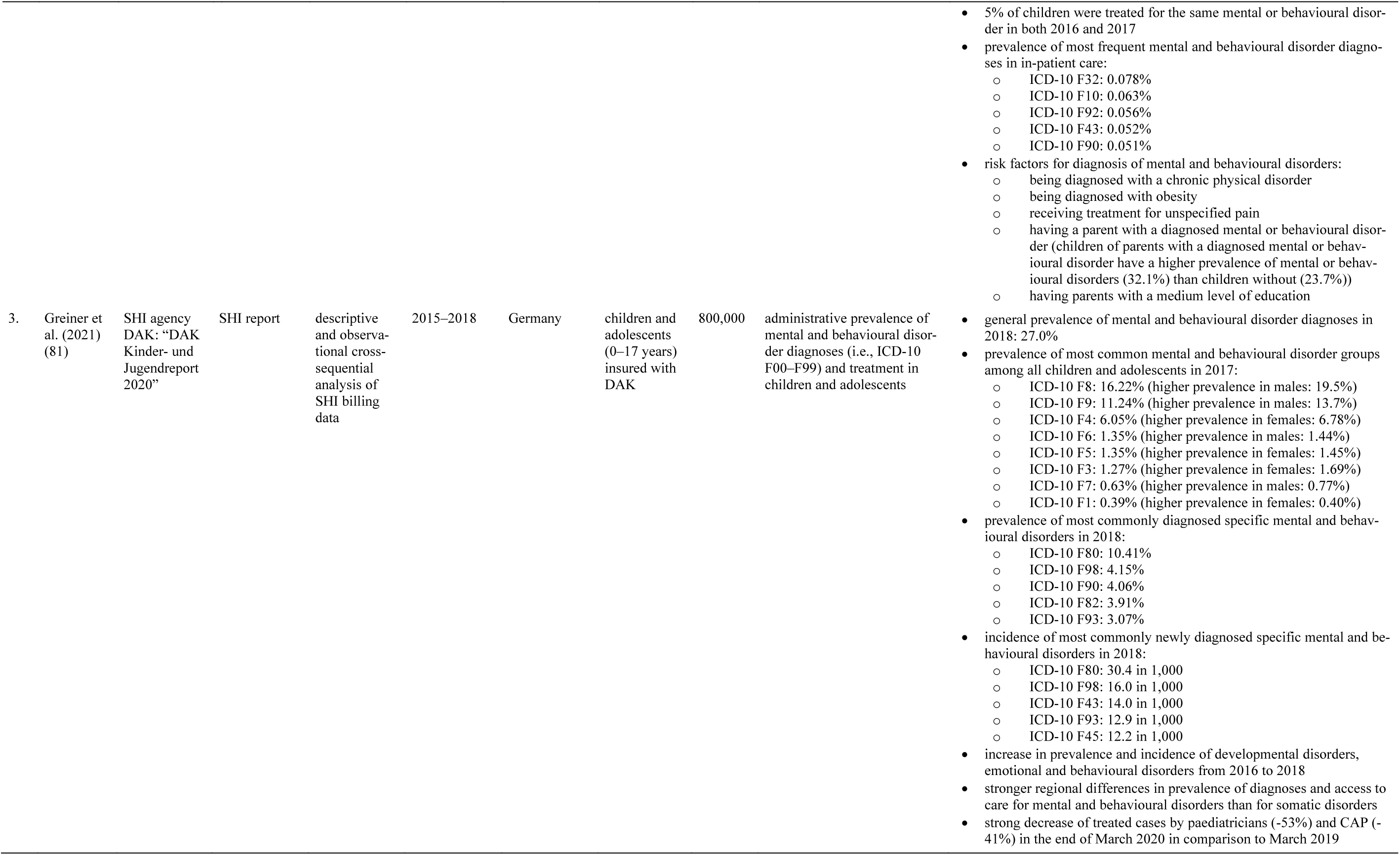
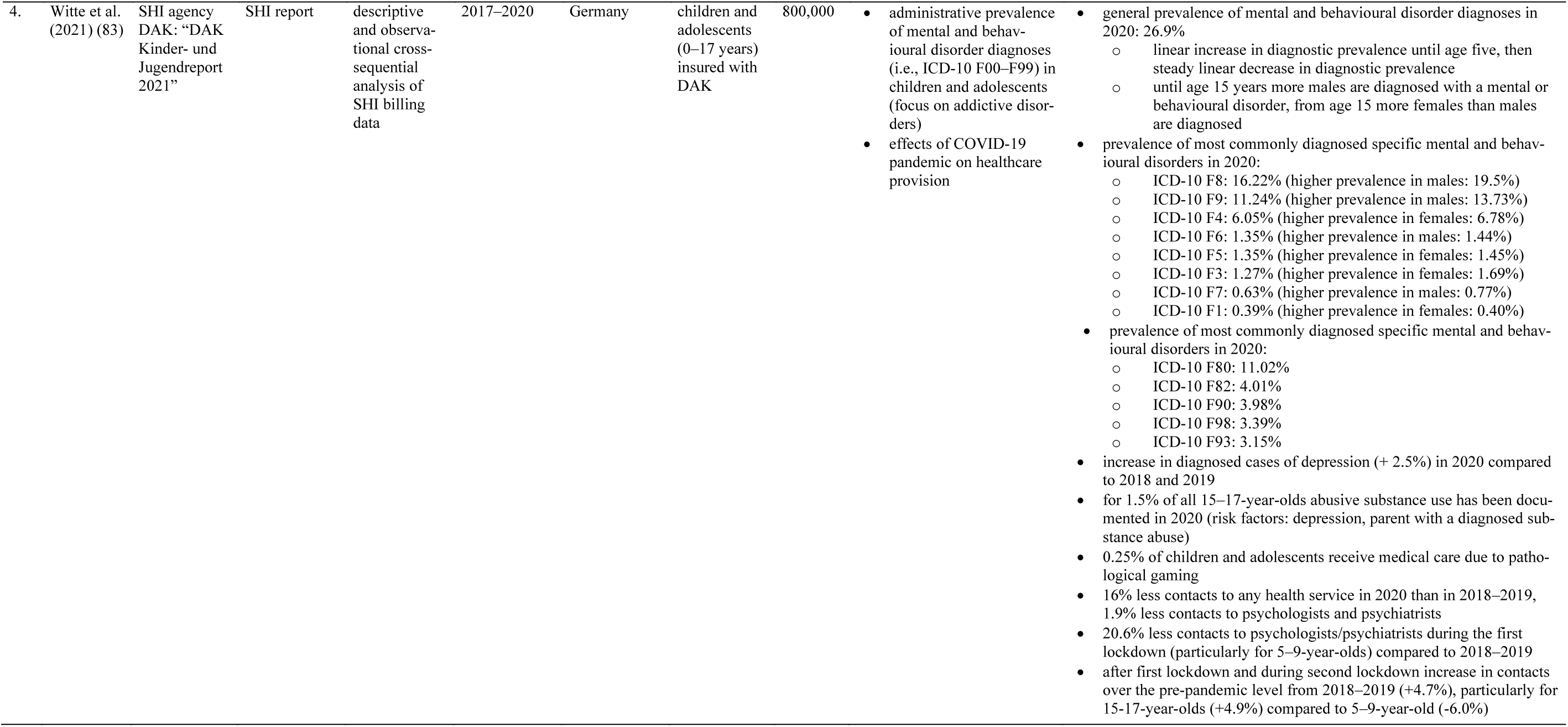
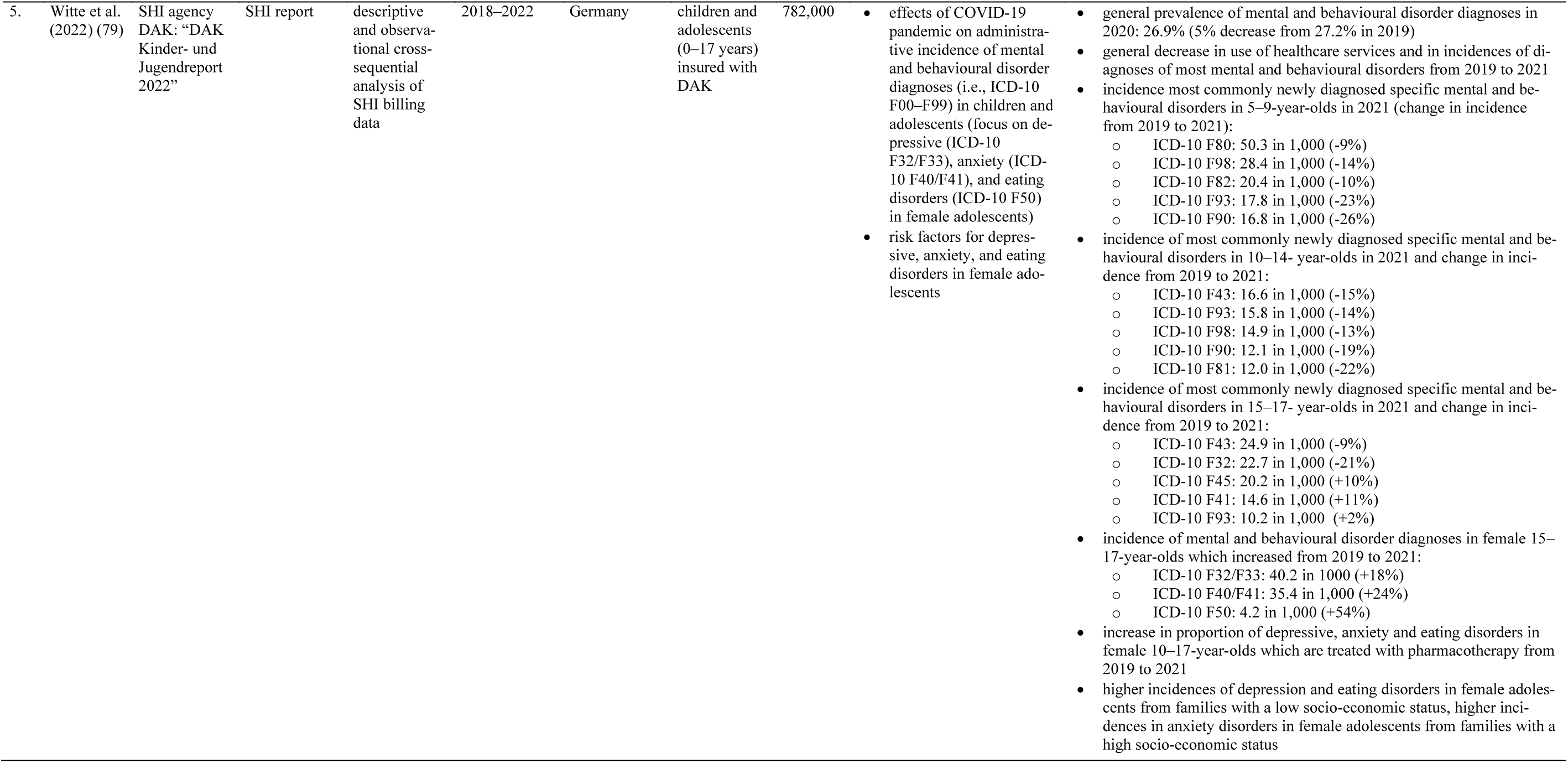
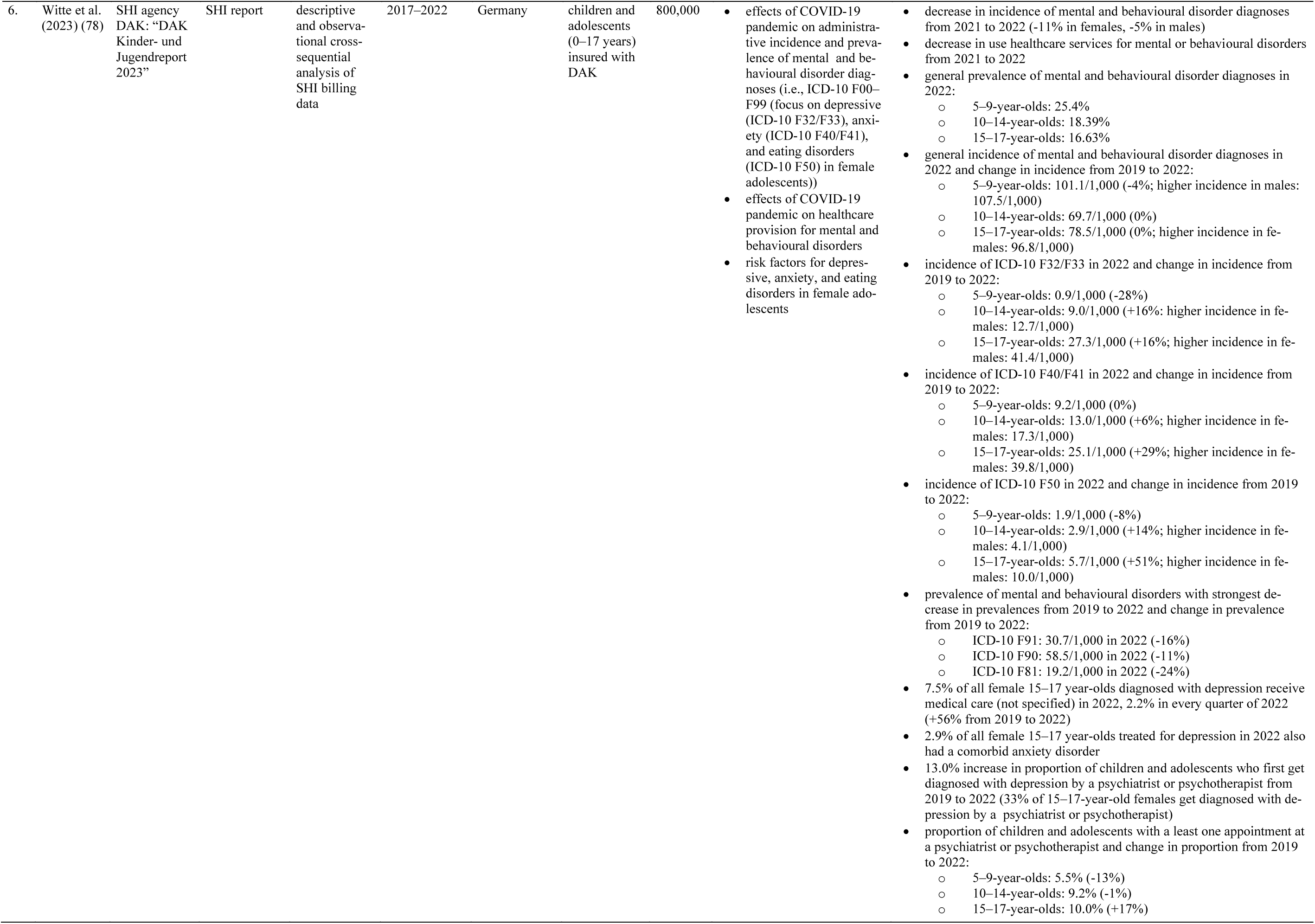
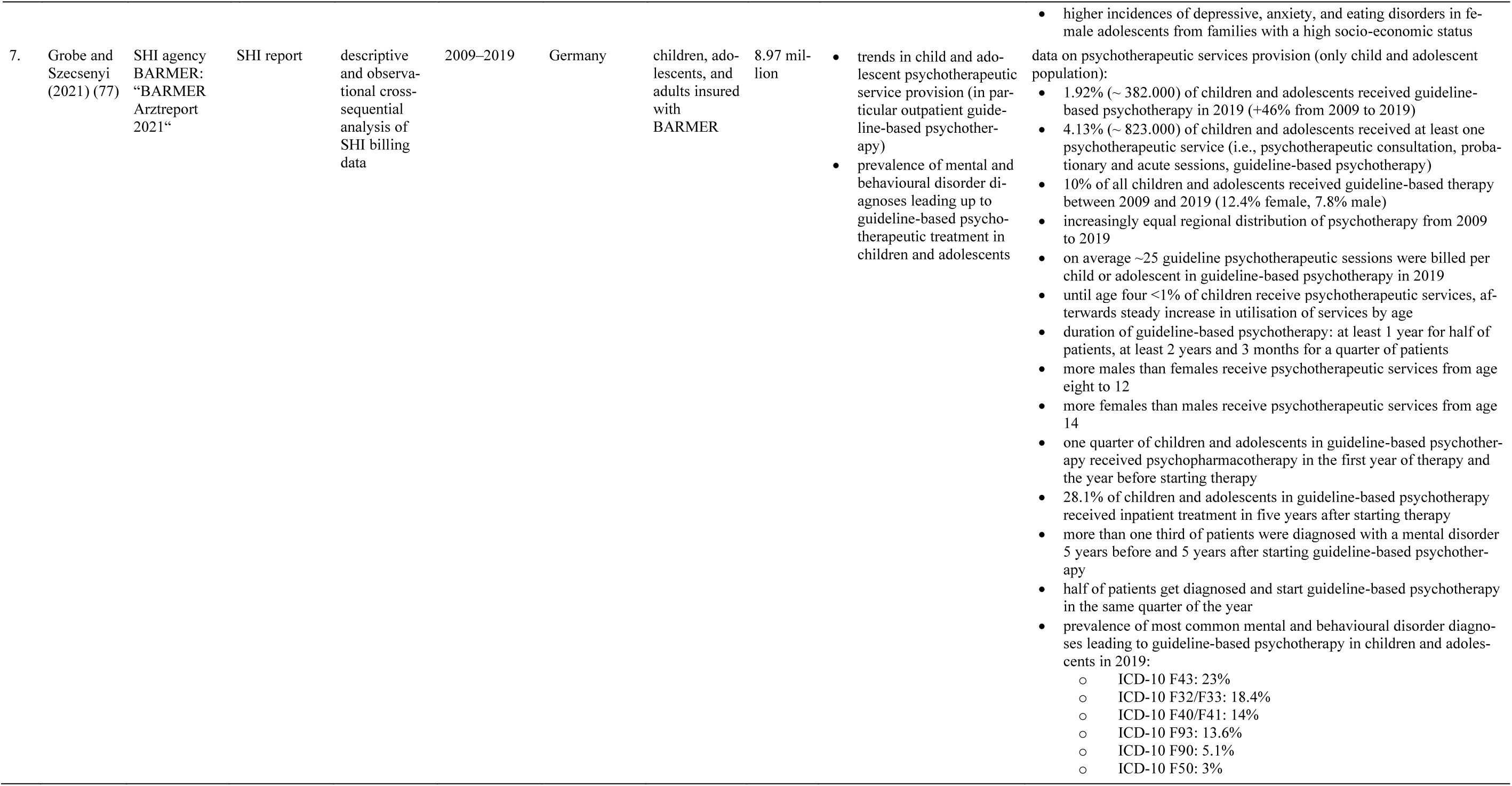
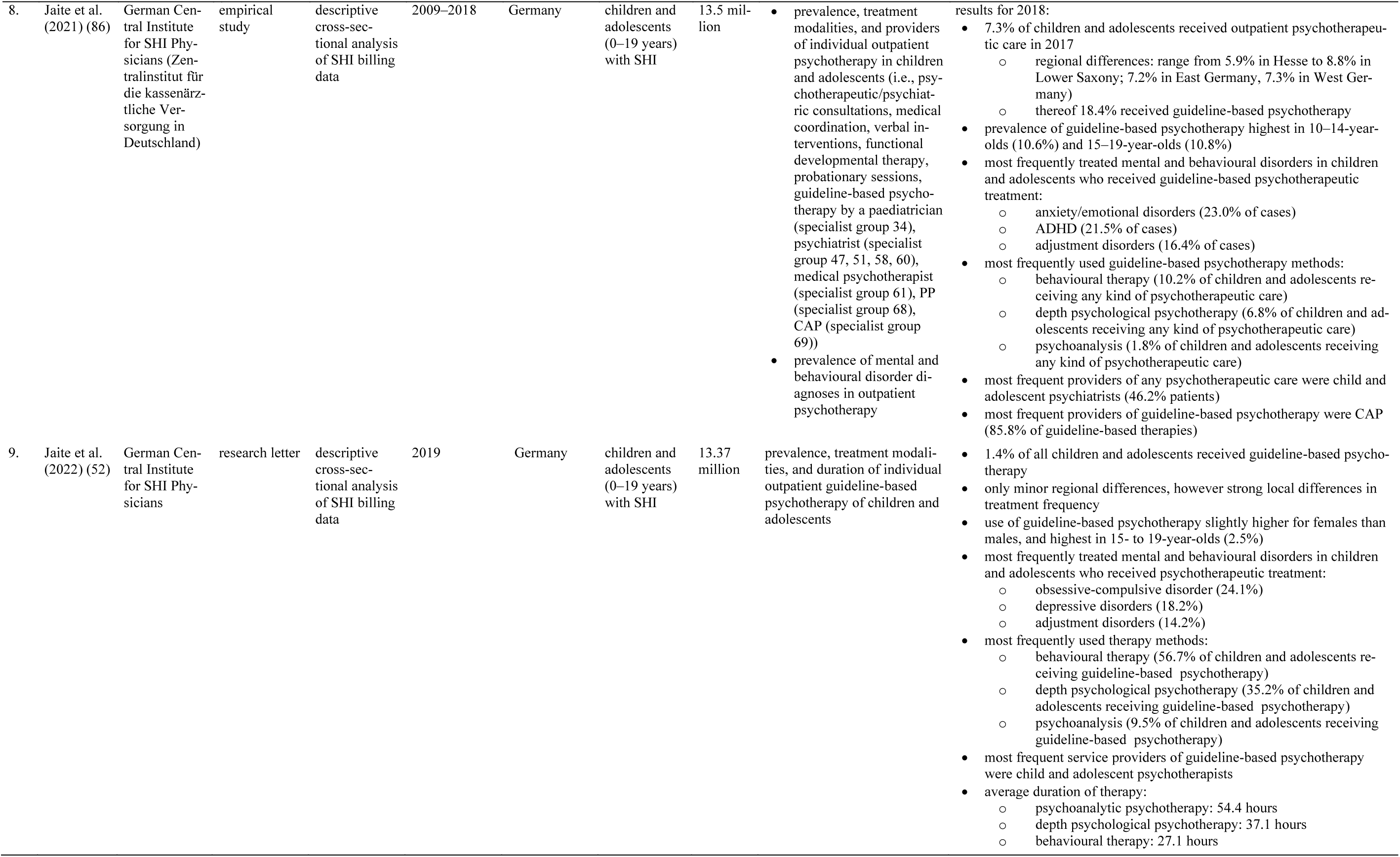
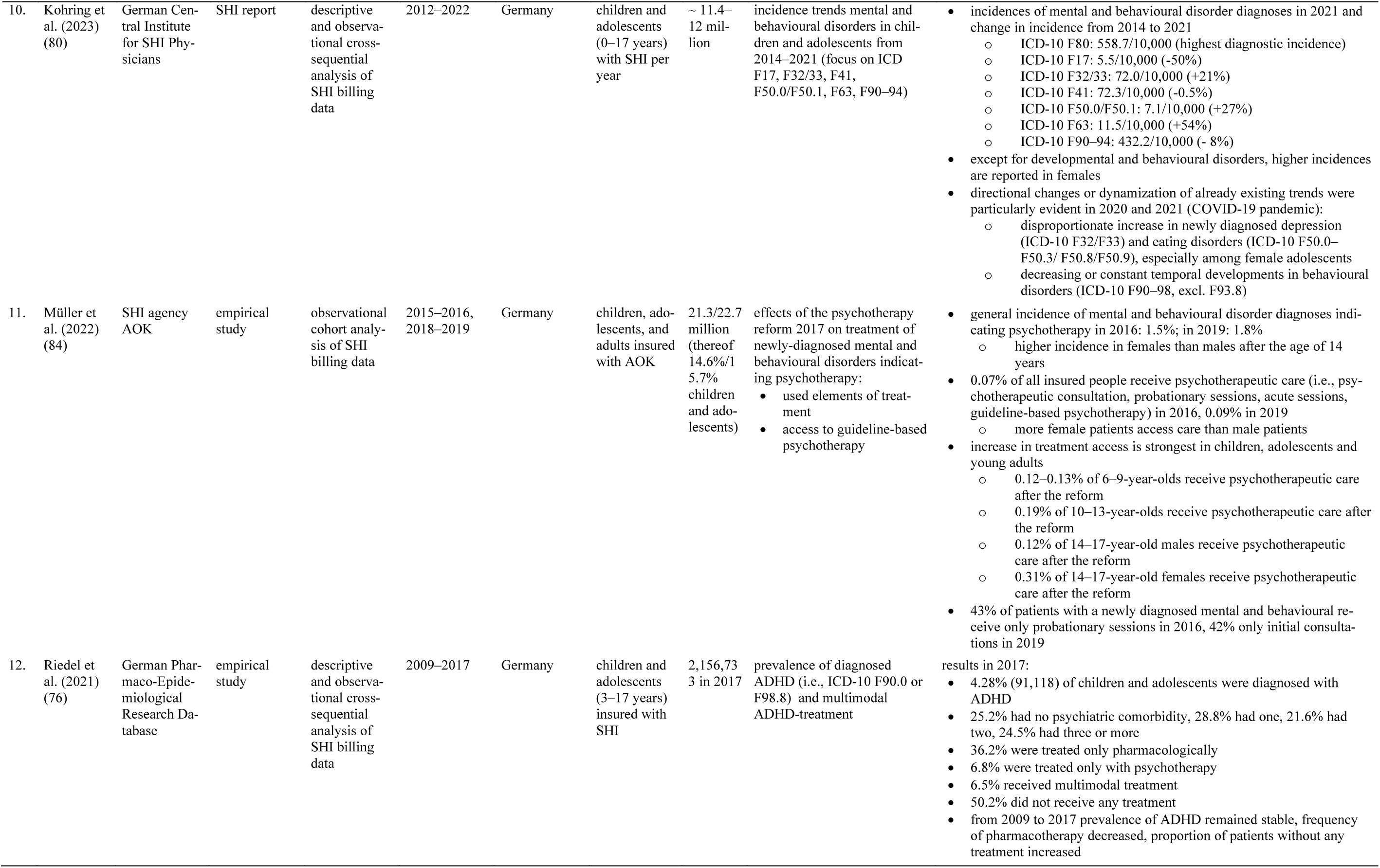
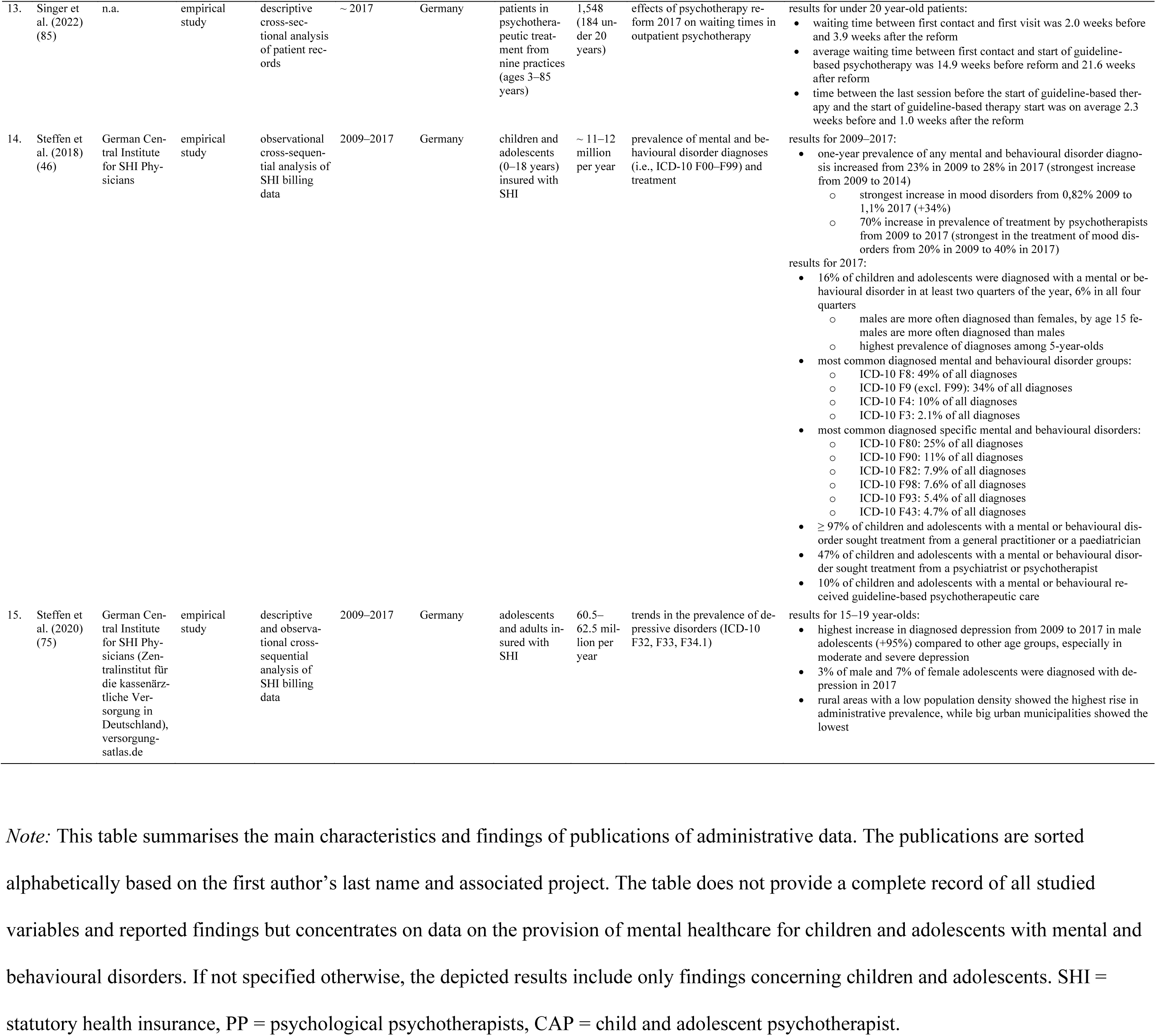
Overview of Administrative Data.

**Table 3.** Overview of Psychotherapist Reports.

| No. | Authors | Association | Type of Article | Method/Design | Time of Assessment | Region | Participants | N | Format | Researched Constructs | Main Findings |
| --- | --- | --- | --- | --- | --- | --- | --- | --- | --- | --- | --- |
| 1. | Bundespsychotherapeutenkammer (2018) (40) | Federal Chamber of Psychotherapists in Germany (Bundespsychotherapeutenkammer (BPtK)) | report of a psychotherapist organisation | descriptive cross-sectional | 11–12/2017 | Germany | SHI-accredited PP<br>SHI-accredited CAP | 9,432 | survey with standardised questionnaire | effects of the reform of the psychotherapy guideline in 2017 on waiting times in outpatient psychotherapy | waiting time for initial psychotherapeutic consultation: 5.7 weeks (CAP: 4.8 weeks)<br>waiting time for acute treatment: 3.1 weeks<br>waiting time for guideline-based psychotherapy: 19.9 weeks (CAP: 17.7 weeks) (particularly long outside of bigger cities and in the Ruhr area)<br>decrease in waiting time for an appointment for an initial psychotherapeutic consultation after reform 2017<br>high variance in waiting time between practices<br>correlation between waiting time and relative amount of psychotherapists in the respective area<br>9.3% of patients receiving an initial psychotherapeutic consultation do not get diagnosed with a mental and behavioural disorder<br>66.3% of psychotherapists offer acute treatment within two weeks, 62.7% said it was difficult to refer patients in need of acute treatment<br>33.5% of psychotherapists reported they could offer 2.6 hours for psychotherapy less per week due to the reform<br>average monthly appointment requests: 22.6, average amount of new patients in 6 months: 13.9 (increase in appointment requests and amount of new patients from 2011 to 2017) |
| 2. | Ostdeutsche Psychotherapeutenkammer (2022) (88–91) | East-German Psychotherapist's Chamber (Ostdeutsche Psychotherapeutenkammer (OPK)) | press statement | descriptive cross-sectional | 03–04/2022 | East Germany | CAP | 206 | survey with standardised questionnaire | effects of the COVID-19 pandemic on mental health and need for mental healthcare of children and adolescents | 91% of CAP said that request for psychotherapy had increased due to the pandemic (63% significant increase, 28% slight increase), increases up to one year, compared to 2–5 months before the pandemic<br>need for psychotherapy increased over 80% for 14–17-year-olds, 30% for 10–13-year-olds<br>CAP reported to offer more initial consultations, increase waiting list capacity, and make appointments far in the future |
| 3. | Psychotherapeutenkammer Niedersachsen (2022) (92) | Psychotherapist's Chamber of Lower Saxony (Psychotherapeutenkammer Niedersachsen (PKN)) | presentation | descriptive cross-sectional | 2021–22 | Lower Saxony, Germany | PP (private practices & SHI-accredited)<br>CAP (private practices & SHI-accredited)<br>inpatient mental healthcare institutions | 1,085, thereof 200 CAP | survey with standardised questionnaire | demand for outpatient psychotherapy (requests, waiting times)<br>patient referral | CAP report to receive 4.7 requests in a week<br>average waiting time for initial consultation 6.3 weeks (median = 4 weeks), similar waiting time for CAP and psychological psychotherapists<br>average waiting time for start of therapy 32 weeks (median = 24 weeks), shorter waiting times for CAP<br>72% of SHI-accredited psychotherapists say that referring patients to other therapists is very difficult or not possible due to colleagues working at full capacity<br>< 2% of SHI-accredited psychotherapists say that they do not experience issues referring patients to other therapists or mental health services |
| 4. | Plötner et al. (2022) (44) | n.a. | empirical study | descriptive cross-sectional | 05/21–07/21 | Germany | CAP | 324 | survey with standardised questionnaire | effects of COVID-19 pandemic on psychotherapeutic care for children and adolescents (waiting times, provided services, mental and behavioural) | • waiting time for initial consultation: 10.2 weeks<br>• waiting time for start of guideline-based psychotherapy: 14.4 weeks<br>• doubling of waiting time for treatment since start of the pandemic<br>• increase in provided sessions per therapist, particularly initial consultations<br>• half of patients experienced pandemic-associated deterioration in mental health<br>• increase in mental disorders since start of the pandemic |
| 5. | Rabe-Menssen et al. (2019) (87) | German Association of Psychotherapists (Deutsche Psychotherapeuten-Vereinigung (DPtV)) | empirical study | descriptive and observational cross-sequential | • 02–04/2017<br>• 02–04/2018 | Germany | members of the DPtV<br>• PP<br>• CAP | 3,018 in 2018, thereof 24% of which treating children and adolescents | survey with standardised questionnaire | waiting time for psychotherapeutic care before and after introduction of obligatory initial consultation | • 44% increase in amount of initial psychotherapeutic consultations from 2017 to 2018<br>• average waiting time for initial consultation decreased from 9.8 in 2017 to 6.5 weeks in 2018 (5.5 weeks for children and adolescents)<br>• average time from request to start of acute treatment: 10.5 weeks (8.8 weeks for children and adolescents), i.e., ~4 weeks after initial consultation → no relevant waiting time<br>• average waiting time for guideline-based psychotherapy increased from 15.3 to 19.6 weeks (18.1 weeks for children and adolescents) due to introduction of obligatory initial consultation → lower threshold to access to initial consultation and diagnostic clarification, but no new treatment capacities<br>• 21% increase in new patients receiving guideline-based therapy, particularly due to acute treatment from 2017 to 2018<br>• 40% of patients having an initial consultation start guideline-based therapy within six months<br>• strong regional variance of waiting times (longest waiting times in Mecklenburg-Western Pomerania, Thuringia, Saarland, Lower Saxony), waiting time is correlated with supply density<br>• reduction of regional differences from 2017 to 2018 |
| 6. | Rabe-Menssen (2021) (41) | German Association of Psychotherapists (DPtV) | empirical study | descriptive and observational cross-sequential | • 01–02/2020<br>• 01–02/2021 | Germany | members of the DPtV<br>• PP<br>• CAP | 4,693 in 2021, thereof 17% CAP | survey with standardised questionnaire | effects of COVID-19 pandemic on:<br>• requests for psychotherapeutic care<br>• waiting time for psychotherapeutic care | • 40.8% increase in weekly requests from 2020 to 2021, on 6.9 requests per week in 2021 (60.3% increase to 5.9 requests for children and adolescents in 2021)<br>• 26% of requests receive appointment for initial consultation (34.5% for children and adolescents)<br>• waiting time for initial consultation: over a month for 49.6% of patients (43.4% for children and adolescents)<br>• waiting time for guideline-based psychotherapy: more than six months for 38.3% of patients (38.8% for children and adolescents)<br>• regional differences: stronger increase in requests in bigger cities, longer waiting times in smaller cities |
| 7. | Rabe-Menssen (2022) (42) | German Association of Psychotherapists (DPtV) | empirical study | descriptive and observational cross-sequential | • 01–02/2020<br>• 01–02/2021<br>• 06/2022 | Germany | members of the DPtV<br>• PP<br>• CAP | 2,270 in 2022, thereof 19% CAP | survey with standardised questionnaire | effects of COVID-19 pandemic on:<br>• requests for psychotherapeutic care<br>• waiting time for psychotherapeutic care | • 40.8% increase in weekly requests from 2020 to 2022, on 6.9 requests per week in 2022 (48% increase to 5.5 requests for children and adolescents in 2022)<br>• 24.6% of requests receive appointment for initial consultation (29.1% for children and adolescents)<br>• waiting time for initial consultation: over a month for 51.0% of patients (similar for children and adolescents)<br>• waiting time for guideline-based psychotherapy: over half a year for 47.4% of patients (similar for children and adolescents)<br>• regional differences: stronger increase in requests in bigger cities, longer waiting times in smaller cities |

*Note:* This table summarises the main characteristics and findings of studies on psychotherapeutic care for children and adolescents from the point of view of psychotherapists. The publications are sorted alphabetically based on the first author's last name. The table does not provide a complete record of all studied variables and reported findings but concentrates on data on the provision of mental healthcare for children and adolescents with mental and behavioural disorders. If not specified otherwise, the depicted results include only findings concerning children and adolescents. PP = psychological psychotherapists, CAP = child and adolescent psychotherapists.

**Table 4.**
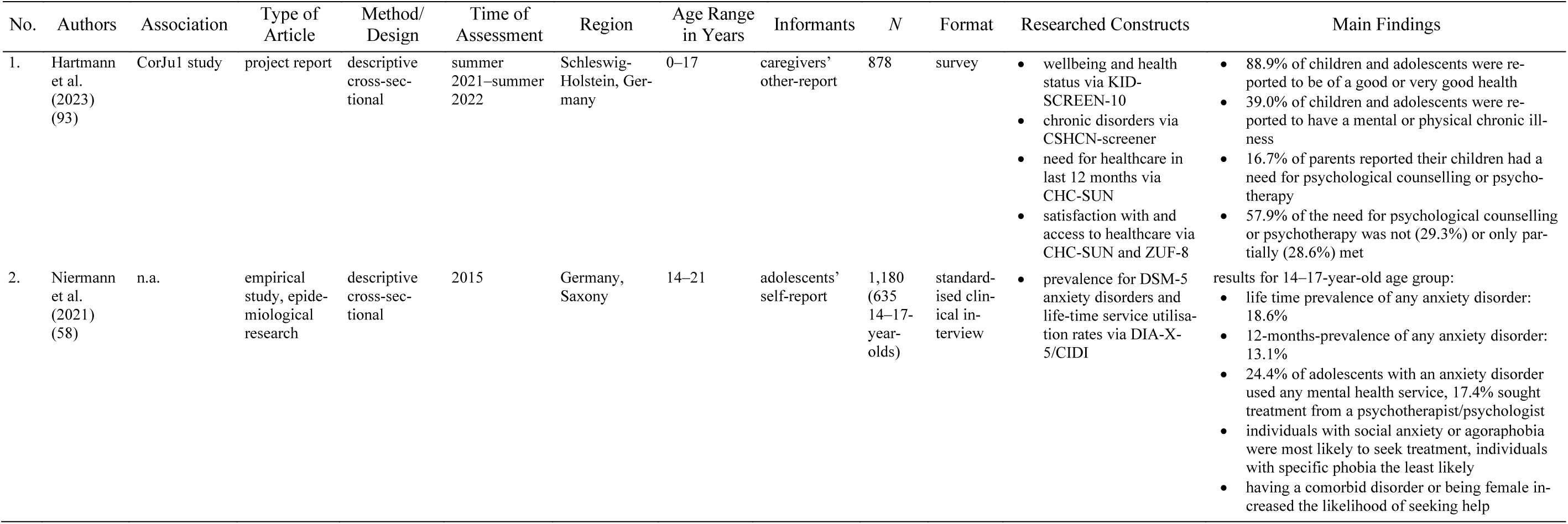
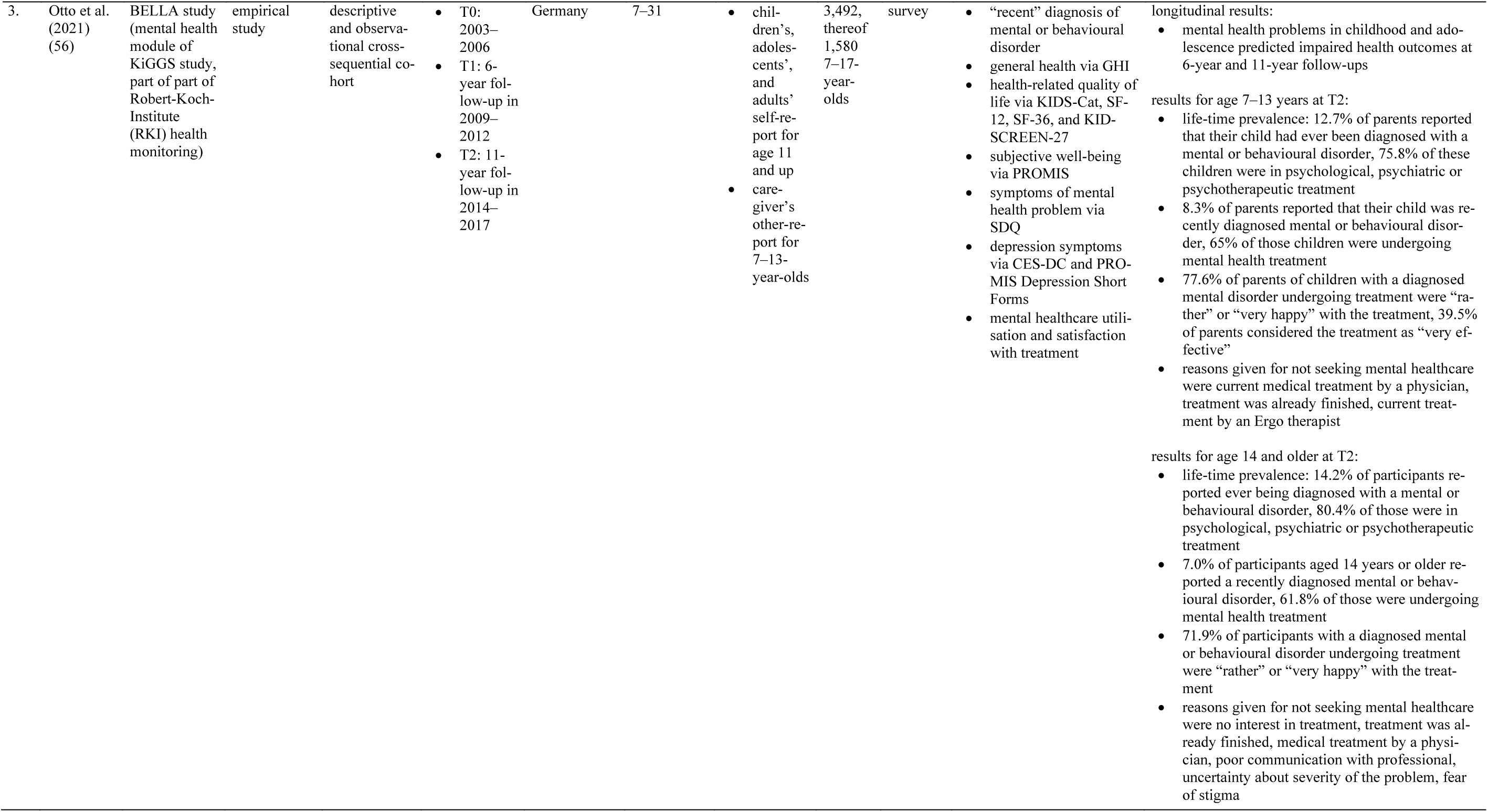
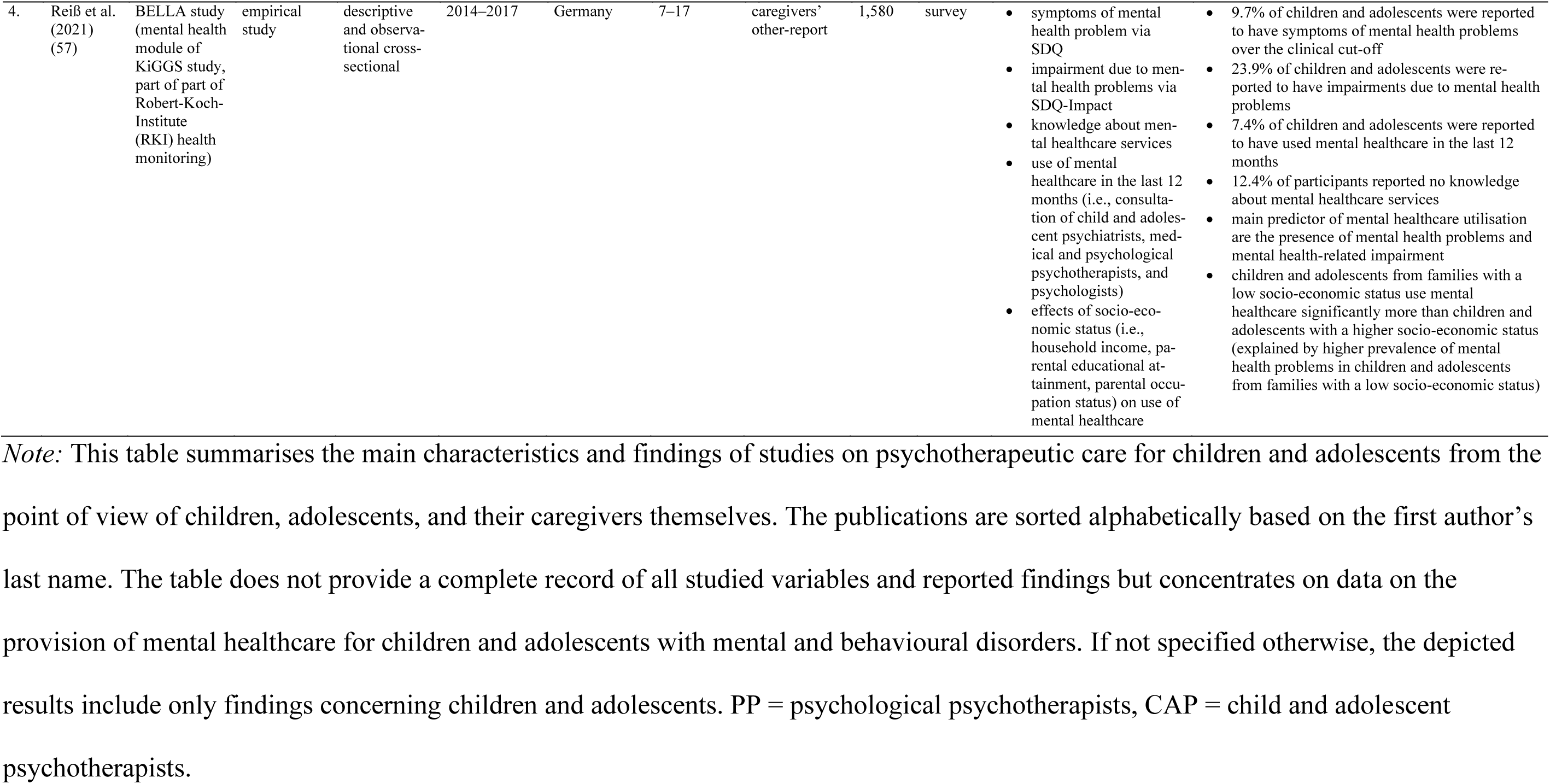
Overview of Patient Reports.

| No. | Authors | Association | Type of Article | Method/Design | Time of Assessment | Region | Age Range in Years | Informants | N | Format | Researched Constructs | Main Findings |
| --- | --- | --- | --- | --- | --- | --- | --- | --- | --- | --- | --- | --- |
| 1. | Hartmann et al. (2023) (93) | CorJu1 study | project report | descriptive cross-sectional | summer 2021–summer 2022 | Schleswig-Holstein, Germany | 0–17 | caregivers’ other-report | 878 | survey | • wellbeing and health status via KID-SCREEN-10<br>• chronic disorders via CSHCN-screener<br>• need for healthcare in last 12 months via CHC-SUN<br>• satisfaction with and access to healthcare via CHC-SUN and ZUF-8 | • 88.9% of children and adolescents were reported to be of a good or very good health<br>• 39.0% of children and adolescents were reported to have a mental or physical chronic illness<br>• 16.7% of parents reported their children had a need for psychological counselling or psychotherapy<br>• 57.9% of the need for psychological counselling or psychotherapy was not (29.3%) or only partially (28.6%) met |
| 2. | Niermann et al. (2021) (58) | n.a. | empirical study, epidemiological research | descriptive cross-sectional | 2015 | Germany, Saxony | 14–21 | adolescents’ self-report | 1,180 (635 14–17-year-olds) | standardised clinical interview | • prevalence for DSM-5 anxiety disorders and life-time service utilisation rates via DIA-X-5/CIDI | results for 14–17-year-old age group:<br>• life time prevalence of any anxiety disorder: 18.6%<br>• 12-months-prevalence of any anxiety disorder: 13.1%<br>• 24.4% of adolescents with an anxiety disorder used any mental health service, 17.4% sought treatment from a psychotherapist/psychologist<br>• individuals with social anxiety or agoraphobia were most likely to seek treatment, individuals with specific phobia the least likely<br>• having a comorbid disorder or being female increased the likelihood of seeking help |

| No. | Authors | Association | Type of Article | Method/ Design | Time of Assessment | Region | Age Range in Years | Informants | N | Format | Researched Constructs | Main Findings |
| --- | --- | --- | --- | --- | --- | --- | --- | --- | --- | --- | --- | --- |
| 3. | Otto et al. (2021) (56) | BELLA study (mental health module of KiGGS study, part of part of Robert-Koch-Institute (RKI) health monitoring) | empirical study | descriptive and observational cross-sequential cohort | T0: 2003–2006<br>T1: 6-year follow-up in 2009–2012<br>T2: 11-year follow-up in 2014–2017 | Germany | 7–31 | children's, adolescents', and adults' self-report for age 11 and up<br>care-giver's other-report for 7–13-year-olds | 3,492, thereof 1,580 7–17-year-olds | survey | “recent” diagnosis of mental or behavioural disorder<br>general health via GHI<br>health-related quality of life via KIDS-Cat, SF-12, SF-36, and KID-SCREEN-27<br>subjective well-being via PROMIS<br>symptoms of mental health problem via SDQ<br>depression symptoms via CES-DC and PROMIS Depression Short Forms<br>mental healthcare utilisation and satisfaction with treatment | longitudinal results:<br>mental health problems in childhood and adolescence predicted impaired health outcomes at 6-year and 11-year follow-ups<br>results for age 7–13 years at T2:<br>life-time prevalence: 12.7% of parents reported that their child had ever been diagnosed with a mental or behavioural disorder, 75.8% of these children were in psychological, psychiatric or psychotherapeutic treatment<br>8.3% of parents reported that their child was recently diagnosed mental or behavioural disorder, 65% of those children were undergoing mental health treatment<br>77.6% of parents of children with a diagnosed mental disorder undergoing treatment were “rather” or “very happy” with the treatment, 39.5% of parents considered the treatment as “very effective”<br>reasons given for not seeking mental healthcare were current medical treatment by a physician, treatment was already finished, current treatment by an Ergo therapist<br>results for age 14 and older at T2:<br>life-time prevalence: 14.2% of participants reported ever being diagnosed with a mental or behavioural disorder, 80.4% of those were in psychological, psychiatric or psychotherapeutic treatment<br>7.0% of participants aged 14 years or older reported a recently diagnosed mental or behavioural disorder, 61.8% of those were undergoing mental health treatment<br>71.9% of participants with a diagnosed mental or behavioural disorder undergoing treatment were “rather” or “very happy” with the treatment<br>reasons given for not seeking mental healthcare were no interest in treatment, treatment was already finished, medical treatment by a physician, poor communication with professional, uncertainty about severity of the problem, fear of stigma |
| 4. | Reiß et al. (2021) (57) | BELLA study (mental health module of KiGGS study, part of part of Robert-Koch-Institute (RKI) health monitoring) | empirical study | descriptive and observational cross-sectional | 2014–2017 | Germany | 7–17 | caregivers' other-report | 1,580 | survey | • symptoms of mental health problem via SDQ<br>• impairment due to mental health problems via SDQ-Impact<br>• knowledge about mental healthcare services<br>• use of mental healthcare in the last 12 months (i.e., consultation of child and adolescent psychiatrists, medical and psychological psychotherapists, and psychologists)<br>• effects of socio-economic status (i.e., household income, parental educational attainment, parental occupation status) on use of mental healthcare | • 9.7% of children and adolescents were reported to have symptoms of mental health problems over the clinical cut-off<br>• 23.9% of children and adolescents were reported to have impairments due to mental health problems<br>• 7.4% of children and adolescents were reported to have used mental healthcare in the last 12 months<br>• 12.4% of participants reported no knowledge about mental healthcare services<br>• main predictor of mental healthcare utilisation are the presence of mental health problems and mental health-related impairment<br>• children and adolescents from families with a low socio-economic status use mental healthcare significantly more than children and adolescents with a higher socio-economic status (explained by higher prevalence of mental health problems in children and adolescents from families with a low socio-economic status) |

*Note:* This table summarises the main characteristics and findings of studies on psychotherapeutic care for children and adolescents from the point of view of children, adolescents, and their caregivers themselves. The publications are sorted alphabetically based on the first author's last name. The table does not provide a complete record of all studied variables and reported findings but concentrates on data on the provision of mental healthcare for children and adolescents with mental and behavioural disorders. If not specified otherwise, the depicted results include only findings concerning children and adolescents. PP = psychological psychotherapists, CAP = child and adolescent psychotherapists.

### 3.2 Epidemiological Studies

Eighteen epidemiological studies on the prevalence of mental or behavioural disorder diagnoses and/or symptoms in German children and adolescents were identified. Ten of these studies had multiple waves of assessment, while eight only had one. Nine studies were last conducted during the COVID-19 pandemic (i.e. between 2020 and 2022) (21, 59–67). The remaining studies were last conducted between 2014 and 2018 (56–58, 68–74). Ten papers are based on the KiGGS-(68, 70), BELLA-(56, 57, 67), or COPSY-study (21, 56, 62–67), which are three interconnected study projects. Therefore these studies partially report on the same samples.

Three studies cover an age range from one to three until 18 years (59, 60, 70). Only two studies concentrate on younger children between one and six years (61, 69), while eight studies include older children and adolescents aged seven to 18 years (21, 57, 62–67), and five studies focus only on adolescents aged 11 to 17 years (58, 68, 71–74).

Only two studies investigate the prevalence of mental or behavioural disorder diagnoses (56, 58), while the rest of the studies investigate a multitude of symptoms of mental or behavioural disorders. One study used in-person structured clinical interviews (58), two studies used standardised telephone interviews (71, 72, 74), and the remaining 15 studies used standardised online or paper-pencil questionnaires to assess symptoms of mental and behavioural disorders. Twelve studies used children’s or adolescents’ self-report (21, 56, 58, 62–68, 71–74), whereas 14 studies used caregiver-reports (21, 56, 57, 59–70).

The detailed prevalence data for the different age groups is presented in Table 5. The data suggests a sharp increase in a variety mental health problems and symptoms of mental and behavioural disorders in the first year of the COVID-19 pandemic, followed by a decrease in prevalence after 2020 (21, 62–67). The latest studies from 2020–2022 suggest that every third to every fourth German child and adolescent suffers from clinically relevant mental health problems (21, 56, 57, 61–67). However, the high variance in methodology, included age groups, and researched outcomes between studies, as well as small numbers of studies investigating the same symptom complex or disorder, do not allow for further conclusions.

**Table 5.** Overview of Epidemiological Prevalence Rates of Mental or Behavioural Disorder Diagnoses and Symptoms.

| Mental or Behavioural Disorder<br>Diagnoses and Symptoms | Pre-COVID-19 Pandemic |  |  | COVID-19 Pandemic |  |  |
| --- | --- | --- | --- | --- | --- | --- |
|  | Younger<br>children | Older children | Adoles-cents | Younger<br>children | Older children | Adoles-cents |
| Mental or Behavioural Disorder<br>Diagnosis | - | 8.3%<br>(56) | 7.0%<br>(56) | - | - | - |
| Mental Health Problems |  | 16.9%<br>(70) |  |  | 55.7–57.4%<br>(59, 60) |  |
|  | - | 9.7%–22%<br>(57, 67) |  | 35.3%<br>(61) | 22.6%–30.9%<br>(21, 57, 62–67) |  |
| Low Health-Related Quality of<br>Life | - | - | - | - | 27.0–47.7%<br>(21, 62–66) |  |
| Depressive Symptoms | - | 11–16%<br>(67) |  | - | 14.2%–24.3%<br>(21, 62–67) |  |
|  | - | - | 8.2%<br>(71, 72) | - | - | - |
| Anxiety Symptoms | 10%<br>(69) | 10–15%<br>(67) |  | - | 24.1%–30.1%<br>(21, 62–67) |  |
| Anxiety Disorder Diagnosis | - | - | 13.1%<br>(58) | - | - | - |
| Eating Disorder Symptoms | - | - | 19.8%<br>(68) | - | - | - |
| Psychosomatic Complaints | - | - | - | - | 44.3%–58.4%<br>(21, 62–66) |  |
| Problem Drinking | - | - | 5%<br>(73) | - | - | - |
| Problematic Internet Use | - | - | 2.6–3.5%<br>(74) | - | - | - |

*Note:* This table summarises the prevalence rates of mental and behavioural disorder diagnoses and symptoms according to epidemiological studies from before (i.e., 2016–2018) and during the COVID-19 pandemic (2020–2022) in younger children (i.e. ~ <6 years), older children (i.e. ~ 6–12 years), and adolescents (i.e. ~ >12 years). ). A dash ("-") indicates that no data was available for the prevalence or incidence of the respective diagnosis.

### 3.3 Administrative Data

We found 15 publications reporting on administrative data on the prevalence and incidence of mental and behavioural disorder diagnoses in children and adolescents in routine practice and on the use of SHI-funded mental healthcare in Germany. Most of these publications reported on broad age ranges from zero to 19 years. Many included data collected over a span of multiple years, five using data from 2009 until 2019 (46, 52, 75–77), three using data from until 2022 (78–80), and the remaining seven being conducted between 2015 and 2020 (47, 52, 81–85).

Eight of these reports were provided by individual German SHI agencies (DAK, BARMER, AOK), with six belonging to the “DAK Gesundheitsreport” series (47, 77–79, 81–84). Five more studies were conducted in association with the German Central Institute for SHI Physicians (46, 52, 75, 80, 86). One study used data from the German Pharmaco-Epidemiological Research Database which combines billing data of four SHI agencies (76). One study analysed records from psychotherapist practices in Germany (85).

#### 3.3.1 Prevalence and Incidence Rates of Mental and Behavioural Disorder Diagnoses

Ten publications report on prevalence or incidence rates of mental or behavioural disorders according to SHI billing data. Most of these papers define a case of a mental or behavioural disorder as the presence of an ICD-10 F-diagnosis (F0–F99), which was at least once coded in the billing data by a practitioner (e.g., paediatrician, psychotherapist, psychiatrist) for a patient in a respective time. The studies consistently report that 26–28% of children were diagnosed with a mental or behavioural disorder both before and during the COVID-19 pandemic (47, 79, 81–83). Around half of these diagnoses are developmental disorders (ICD-10 F8), one third are behavioural and emotional disorders with onset in childhood and adolescence (ICD-10 F9), 10% are anxiety, dissociative, stress-related, somatoform, and other nonpsychotic mental disorders (ICD-10 F4), and 2% mood disorders (ICD-10 F3) (46, 47, 81–83). The prevalence and incidence of specific ICD-10 diagnoses before and during the COVID-19 pandemic are displayed in Table 6.

**Table 6.** Overview of Administrative Prevalence Rates of Mental or Behavioural Disorder Diagnoses in SHI Billing Data.

| Diagnosis<br>(According to<br>ICD-10) | One-Year Prevalence |  | One-Year Incidence (in 1,000) |  |
| --- | --- | --- | --- | --- |
|  | Pre-COVID-19 Pandemic | COVID-19 Pandemic | Pre-COVID-19<br>Pandemic | COVID-19 Pandemic |
| Any F-diagnosis | 25.9%–28% (47, 81, 82) | 26.9% (79, 83)<br>25.4% of 5–9-y.-o.<br>(78)<br>18.39% of 10–14-y.-<br>o. (78)<br>16.63% of 15–17-y.-<br>o. (78) | - | - |
| F1 | 0.31%–0.39% (47, 81, 82) | 0.39% (83) | - | - |
| F17 | - | - | - | 0.55 (80) |
| F2 | 0.05% (47, 81, 82) | 0,05% (83) |  |  |
| F3 | 1.08%–1.28% (47, 81, 82) | 1.27% (83) |  |  |
| F32 & 33 | 1.5% (82)<br>3% of 15–19-y.-o. males (82)<br>7% of 15–19-y.-o. females<br>(82) | - | - | 7.2 (80)<br>0.9 5–9-y.-o. (78)<br>9.0 10–14-y.-o. (78)<br>22.7 (only F32) –27.3 15–17-y.-<br>o. (78, 79) |
| F4 | 5.28%–6.05% (47, 81, 82) | 6.05% (83) | - | - |
| F40 & 41 | 0.67% (only F40), 1.29% (only<br>F41) (82) | - | - | 7.2 (only F41) (80)<br>9.2 5–9-y.-o. (78)<br>13.0 10–14-y.-o. (78)<br>25.1 15–17-y.-o. (78, 79) |
| F42 | 0.24% (82) | - | - | - |
| F43 | 3.47% (82) | - | 14.0 (81) | 16.6 10–14-y.-o. (79)<br>24.9 15–17-y.-o. (79) |
| F45 | 2.64% (82) | - | 12.2 (81) | 20.2 15–17-y.-o. (79) |
|  | Pre-COVID-19 Pandemic | COVID-19 Pandemic | Pre-COVID-19<br>Pandemic | COVID-19 Pandemic |
| F5 | 1.24%–1.38% (47, 81, 82) | 1.35% (83) | - | - |
| F50 | - | - | - | 0.7 (only F50.0, F50.1) (80)<br>1.9 5–9-y.-o. (78)<br>2.9 10–14-y.-o. (78)<br>5.7 15–17-y.-o. (78) |
| F6 | 1.28%–1.35% (47, 81, 82) | 1.35% (83) | - | - |
| F63 | - | - | - | 1.15 (80) |
| F7 | 0.63%–0.65% (47, 81, 82) | 0.63% (83) | - | - |
| F8 | 14.83%–16.22% (47, 81, 82) | 16.22% (83) | - | - |
| F80 | 9.52%–10.41% (47, 81, 82) | 11.02% (83) | 30.4 (81) | 55.9 (80)<br>50.3 5–9-y.-o. (79) |
| F81 | - | - | - | 12.0 10–14-y.-o. (79) |
| F82 | 3.49%–3.91% (47, 81, 82) | 4.01% (83) | - | 20.4 5–9-y.-o. (79) |
| F9 | 10.5%–11.24% (47, 81, 82) | 11.24% (83) | - | - |
| F90 | 3.88%–4.28% (47, 81, 82) | 3.98% (83) | - | 16.8 5–9-y.-o. (79)<br>12.1 10–14-y.-o. (79) |
| F93 | 2.68%–3.07% (47, 81, 82) | 3.15% (83) | 12.9 (81) | 17.8 5–9-y.-o. (79)<br>15.8 10–14-y.-o. (79)<br>10.2 15–17-y.-o. (79) |
| F98 | 3.71%–4.15% (47, 81, 82) | 3.39% (83) | 16.0 (81) | 28.4 5–9-y.-o. (79)<br>14.9 10–14-y.-o. (79) |

*Note:* This table summarizes the administrative one-year prevalence and incidence rates of mental and behavioural disorder diagnoses in children and adolescents (0–18 years unless specified otherwise) according to the ICD-10 in SHI billing data from the period before (2016–2018) and during the COVID-19 pandemic (2020–2022). A dash (“-”) indicates that no data was available for the prevalence or incidence of the respective diagnosis. y.-o. = year-olds.

The publications indicate a small but steady increase of mental and behavioural disorder diagnoses before the COVID-19 pandemic, which was followed by a general decrease in use of healthcare services and incidence numbers of most mental and behavioural disorders during the pandemic (78, 79, 81). However, while the incidence of behavioural disorders seemed to stay constant or decrease, multiple papers report a disproportionate increase in incidence numbers of depressive, anxiety, and eating disorders among female adolescents during the pandemic (78–80).

Over the lifespan, data conclusively shows a linear increase in prevalence of mental or behavioural disorders until age five, followed by a steady linear decrease in prevalence (46, 47, 82). While male children and adolescents are more often diagnosed with a mental or behavioural disorder than female children and adolescents until age 15, from age 15 more female than male adolescents are diagnosed (46, 47, 80, 82–84). In childhood, mental and behavioural disorder diagnoses are dominated by developmental and behavioural disorders (ICD-10 F8 & F9); in adolescence the most prevalent disorders are mood and anxiety disorders (ICD-10 F3 & F4) (53, 58, 59, 61–63). Specifically, the highest prevalence rates of developmental and behavioural disorders (ICD-10 F8 & F9) are found in male children, whereas prevalence rates of substance use, anxiety, eating, and personality disorders (ICD-10 F1, F3, F4, F5 & F6) are highest among female adolescents (46, 47, 80, 82–84).

#### 3.3.2 Use and Provision of Statutory Health Insurance Funded Mental Healthcare

Twelve publications provide data on access and use of SHI-funded outpatient mental healthcare (i.e., any medical or psychological treatment of a mental or behavioural disorder provided by a physician, psychologist, or psychotherapist), some of them also more specifically on psychotherapeutic care. However, the papers differ greatly in their definitions of psychotherapeutic services/psychotherapeutic care, which leads to variance in the provided data.

Nonetheless, the data conclusively shows that there is a general increase in the use of psychotherapeutic care from age five: While less than 1% of under 5-year-olds are in psychotherapeutic care, around 6% of 5–9-year-olds and 11% of 10–19-year-olds receive psychotherapeutic treatment (52, 77, 82, 84, 86). Consistent with gender differences in prevalence of mental and behavioural disorders, more male than female children use psychotherapeutic care in late childhood, whereas more female than male adolescents use receive psychotherapeutic care in adolescence (46, 77, 82, 84).

In the years shortly before the COVID-19 pandemic (i.e. 2017–2019) the publications indicate that between 5 to 7% of all children and adolescents used outpatient mental healthcare services in the broader sense (i.e., psychotherapeutic/psychiatric consultations, medical coordination, verbal interventions, functional developmental therapy, probationary sessions, guideline-based psychotherapy provided by a paediatrician, psychiatrist, medical, psychological, or child and adolescent psychotherapist) (82, 86). 0.2–2% of all children and adolescents were reported to receive psychotherapeutic care in the narrower sense in a year (i.e., guideline-based psychotherapy provided by a medical, psychological, or child and adolescent psychotherapist) (52, 77, 84). Over a span of ten years, 10% of all children and adolescents were recorded to have accessed guideline-based psychotherapy (46, 77). Of those children and adolescents who were newly diagnosed with a mental or behavioural disorder, nearly half received only initial consultations and no further treatment, and only one in ten of those children and adolescents received guideline-based psychotherapy (46, 84).

Data further reveals that 97% of children and adolescents with a mental or behavioural disorder sought treatment from a general practitioner or a paediatrician, while only half sought treatment from a psychiatrist or psychotherapist (46). However, the prevalence of cases treated by psychotherapists increased over the years before the COVID-19 pandemic (84). Most patients receiving psychotherapeutic care in the broader sense were treated by a psychiatrist, whereas the most frequent providers of guideline-based psychotherapy were child and adolescent psychotherapists (52, 86).

Information on waiting times between the initial consultation and the start of guideline-based psychotherapy is scarce in the included publications but indicates that half of patients got initially diagnosed and started guideline psychotherapy in the same quarter of the year, while on average patients had to wait 22 weeks between the initial consultation and the start of guideline-based psychotherapy (77, 85).

Diagnoses most frequently treated in psychotherapeutic outpatient care included anxiety, compulsive and stress-related disorders (ICD-10 F40, F41, F42 & F43), depressive disorders (ICD-10 F32 & F33), hyperkinetic disorders (ICD-10 F90), and emotional disorders with onset in childhood (ICD-10 F93) (52, 77, 82, 86). However, there is strong variance in the reported proportions of these diagnoses in outpatient psychotherapeutic care between different publications.

During the COVID-19 pandemic, the reports recorded a general decrease in use of healthcare services, including visits to paediatricians, psychiatrists, and psychotherapists, particularly during the first lockdown in spring 2020 (79, 82, 83). Consequently, administrative incidence rates of most mental and behavioural diagnoses initially dropped. After the first lockdown the amount of contacts surpassed the pre-pandemic level (83). Two years into the pandemic, data shows that significantly less younger children and significantly more adolescents receive outpatient psychotherapeutic care than before the pandemic (78).

### 3.4 Psychotherapists’ Reports

Seven studies assessed the state of psychotherapeutic care for children and adolescents, particularly waiting times and provided services, from the perspective of psychotherapists. Two of these studies were conducted in 2017 to 2018 during the introduction of the obligatory initial psychotherapeutic consultation. The rest of the studies were conducted between 2020 and 2022.

It was shown that the introduction of the obligatory initial consultation in 2017 led to an increase in patients accessing care, however, at the same time also to an increase of waiting time for guideline-based psychotherapy (40, 87). During the pandemic, psychotherapists consistently reported a growing demand and need for psychotherapy due to pandemic-related increases in mental disorders and deterioration of existing mental health problems (44, 88–91).

The studies concordantly report that psychotherapists received five to six requests for an initial appointment per week, of which only one third could be offered an appointment during the pandemic (41, 42, 44, 87, 92). Waiting times for an initial psychotherapeutic consultation seem to have increased from approximately five to six weeks before the pandemic to six to ten weeks during the pandemic (40–42, 44, 87, 92). After the initial consultation, patients had to wait another four months before the pandemic and three to seven months during the pandemic to start guideline-based psychotherapy (40–42, 44, 87, 92). Roughly every second child or adolescent had to wait over a month for a consultation after receiving an appointment and more than six months for the start of guideline-based psychotherapy during the pandemic (41, 42). However, multiple studies found high variance in waiting times between individual practices and between regions (40–42, 87). Furthermore, psychotherapists stated that they found it very difficult or even impossible to refer patients to other therapists before and during the pandemic (40, 92).

### 3.5 Patients’ Reports

Four included studies investigated the provision psychotherapeutic care for children and adolescents from the perspective of the patients’ or their families themselves. Three of these studies were conducted between 2014 to 2017 before the introduction of the obligatory initial psychotherapeutic consultation, while one study was conducted during the COVID-19 pandemic in 2021 and 2022.

The studies show that approximately 7% of the general population children and adolescents reported using mental healthcare in the broader sense (i.e., at least one consultation at a psychiatrist, psychotherapist or psychologist) before the pandemic (57). In a sample of children and adolescents with a diagnosed mental or behavioural disorder, about two thirds to three quarters of those children and adolescents were reported to received such care (56). During the COVID-19 pandemic, circa 17% of caregivers stated that their child was in need of mental healthcare, while this need was not or only partially met in the majority of cases (93). Reasons for the discrepancy between the perceived and actual need for psychotherapeutic treatment and the care received include a lack of awareness about mental healthcare services, treatment by other professionals, fear of stigma, and other factors. Socioeconomic status, the severity of mental health problems and related impairment, the specific diagnosis and comorbidities, and gender moderated the likelihood of seeking help for mental health problems (57, 58, 93).

## 4 Discussion

Worldwide, increasing numbers of children and adolescents are affected by mental or behavioural disorders requiring psychotherapeutic treatment, but often face barriers to accessing outpatient psychotherapeutic care. Here, we present a scoping literature review on the state of outpatient psychotherapeutic care for children and adolescents in Germany and its assessment. In Germany, mental healthcare is predominantly funded by statutory health insurance and offers high treatment capacities in the inpatient sector, but reportedly lacks sufficient treatment capacities in the outpatient sector in comparison to other high-resource countries. Comprehensive, systematic, and multimodal analyses of psychotherapeutic care provision, necessary for empirically-based demand planning, are lacking.

This review was conducted according to PRISMA-ScR guidelines (54). We included 41 papers based on epidemiological studies, administrative data, and provider and patient reports and published between April 2018 and November 2023. Our assessment focussed on the prevalence and incidence rates of mental and behavioural disorder symptoms and diagnoses, as well as the demand and need for, the provision and use, and the availability and accessibility of outpatient psychotherapeutic care in German children and adolescents. Additionally, we examined the suitability of different data sources for adequately assessing outpatient psychotherapeutic care.

### 4.1 Prevalence of Mental and Behavioural Disorder Diagnoses and Symptoms in German Children and Adolescents

Based on epidemiological studies, we aimed to address the first research question concerning the prevalence of mental and behavioural disorder symptoms and diagnoses in children and adolescents in Germany in recent years. Unfortunately, we found very few studies using systematic and standardized clinical interviews to estimate disorder prevalence rates. There seems to be a lack of epidemiological data on mental and behavioural disorders in German children and adolescents, in particular in younger children. Older national and international reviews and meta-analyses estimated the prevalence of mental and behavioural disorders in childhood and adolescence to range between 10–25%, with significant variance in the time of assessment, case definition, and methodology (1–3, 94, 95). A promising and more recent Austrian study from 2016 assessed the prevalence of mental and behavioural disorders according to the DSM-5 in 10–18-year-olds with both clinical questionnaires and a structured interview and shows that 23.9% of children and adolescents met the criteria of a mental or behavioural disorder (96). When comparing these studies, it is important to note that only one exclusively reported on children and adolescents in Germany (94). Additionally, some of the reviews include data dating back to the 1970s, with one study even reaching back to the 1950s. Although epidemiological estimates are generally considered to be more stable and less time-sensitive, it is likely that the prevalence rates of mental and behavioural disorders in children and adolescents have changed over the last decades due to shifts in their environments (e.g., political and societal developments, exposure to global crises, digitalization). Moreover, the conceptualization and diagnostic criteria for mental and behavioural disorders have evolved over time, which is likely to influence epidemiological data. However, the impact of these changes cannot be estimated based on the current research, as recent epidemiological data on German children and adolescents is lacking.

Nevertheless, we found several studies using clinical questionnaires to assess the prevalence of self- or caregiver-reported mental health problems or psychopathological symptoms as an estimate of child and adolescent mental health, especially during the COVID-19 pandemic. This data suggests a general increase in mental health problems from before the pandemic to during the pandemic, followed by a slow decrease after the initial year of the pandemic in 2020 (21, 57, 62–67). Studies conducted between 2020 and 2022 indicate that at least every third to every fourth child and adolescent was affected by mental health problems during the pandemic (21, 62–67). However, the high variance in methods, researched constructs, and included age groups limits the synthesis of these studies’ results. Questionnaire data alone also does not allow for conclusions on disorder prevalence rates. Furthermore, studies on the mental health of children and adolescents conducted after 2022 are still lacking.

Consequently, the first research question cannot be sufficiently answered in light of the current database. This underscores the urgent need for standardized, systematic, and longitudinal assessments of child and adolescent mental health in Germany beyond mere questionnaire measures.

### 4.2 Current State of Outpatient Psychotherapy

We attempted to evaluate the current state of outpatient psychotherapeutic care by examining both epidemiological and administrative data, alongside reports from psychotherapists, as well as children, adolescents, and their families as potential patients. Specifically, we aimed to estimate the need and demand for, the use and provision of, and the accessibility and availability of psychotherapeutic care to address our second research question.

#### 4.2.1 Need and Demand for Psychotherapeutic Care

As discussed above, there is a lack of studies estimating the prevalence of mental and behavioural disorders indicating psychotherapeutic treatment in children and adolescents in Germany. Consequently, it is difficult to estimate the objective need for psychotherapeutic treatment based on recent epidemiological data.

Recent data from the statutory healthcare system indicate that the prevalence rates of mental and behavioural disorders among children and adolescents are slightly higher than those reported in older epidemiological studies, now ranging between 26–28% (47, 79, 81–83). However, these administrative numbers do not reflect the objective need for psychotherapy but merely the number of billed diagnoses in routine care, which usually have lower validity than diagnoses derived from epidemiological studies. Although most mental and behavioural disorders can be treated with psychotherapy, certain disorders are not primarily an indication for psychotherapy, e.g., intellectual disability (ICD-10 F7) or developmental disorders (ICD-10 F8). As these disorders are usually included in the general administrative estimate of mental and behavioural disorders, these estimates do not equal the need for psychotherapy. In fact, roughly half of all mental and behavioural disorder diagnoses in routine care are developmental disorders (46, 47, 81–83). Thus, according to administrative data, it can be cautiously estimated that only 13–14% of all children and adolescents have a diagnoses a mental or behavioural disorder that can be primarily treated with psychotherapy (e.g., emotional and behavioural disorders with onset in childhood and adolescence (ICD-10 F9), anxiety, compulsive and stress-related disorders (ICD-10 F4), mood disorders (ICD-10 F3)). However, this number does not include children and adolescents who might fulfil the criteria of a mental or behavioural disorder but have not yet been diagnosed in routine care. At the same time, it may include a significant number of false-positive diagnoses, as the diagnostic quality is not accounted for in routine data, leading to concerns about the validity of these diagnoses.

Regarding the subjective need or demand for psychotherapy, we again did not find many studies examining children’s and adolescents’ self-assessed or caregiver-assessed need or wish for treatment. One study reported that during the COVID-19 pandemic, around 17% of caregivers stated that their child was in need of mental healthcare and that this need was not or only partially met in the majority of cases (93). Taken together with the data from epidemiological and administrative data, it can be cautiously summarised that roughly every sixth to seventh child and adolescent in Germany is in need of psychotherapeutic treatment. This equates to approximately three million children and adolescents (97).

#### 4.2.2 Use and Provision of Psychotherapeutic Care

It is important to note that the publications we reviewed vary in their definitions of mental or psychotherapeutic healthcare and frequently do not provide data specifically on psychotherapeutic care. These inconsistencies contribute to significant variance in data and, at times, result in contradictory findings.

Administrative data suggests that nearly all families with children and adolescents with a mental or behavioural disorder initially consult a paediatrician or general practitioner regarding the child’s or adolescent’s mental health problems (46). Only half of them come into contact with a mental health specialist, e.g., a psychiatrist or psychotherapist (46). Other publications suggest that only 5–7% of all children and adolescents, and therefore only a quarter to a third of children and adolescents with a mental or behavioural disorder, receive mental healthcare in the broader sense (82, 86). This includes services provided not only by a medical, psychological, or child and adolescent psychotherapist but also by a general practitioner, paediatrician, or psychiatrist in connection to a mental or behavioural disorder diagnosis. Only 0.2–2% of all children and adolescents receive guideline-based psychotherapy, translating to up to 10% of children and adolescents with a mental or behavioural disorder (52, 77, 84).

Regarding age and gender differences in the use of mental healthcare, administrative data suggests a peak in diagnosis prevalence at age five, as most developmental disorders are diagnosed at this age (46, 47, 82). Until age 15, more male than female children and adolescents are diagnosed; afterwards, more female than male adolescents receive a diagnosis (46, 47, 80, 82–84). This aligns with gender differences in the type of diagnosis: while male children and adolescents are more frequently diagnosed with developmental and behavioural disorders, typically occurring first in early childhood, female children and adolescents are more often diagnosed with mood, anxiety, eating, personality, and substance abuse disorders, which typically only start in late childhood or adolescence (46, 47, 80, 82–84). These trends are also reflected in age and gender differences in the use of mental healthcare (46, 77, 82, 84). However, psychotherapy is mostly used by older children and adolescents and less by younger children (52, 77, 82, 84, 86).

Considering changes over time, it seems that an increasing number of children and adolescents are treated by psychotherapists, while fewer cases are only treated by a paediatrician or general practitioner (46). This coincides with the introduction of the obligatory initial consultation in 2017, which every SHI-accredited psychotherapist in private practice is required to provide. This initial consultation serves as a preliminary diagnostic assessment and treatment recommendation, as well as a first step in accessing psychotherapeutic care. However, an appointment for an initial consultation often does not result in a therapy place at the respective practice. Data shows that due to this reform, significantly more patients access psychotherapeutic care, while waiting times for guideline-based psychotherapy further increase (40, 85, 87). This is because the required number of initial consultations a practice must provide reduce the capacity for guideline-based psychotherapy sessions. The COVID-19 pandemic put a heavy strain on the healthcare system in general and therefore also affected the provision of psychotherapeutic care. Administrative data indicates a general decrease in psychotherapeutic service provision and administrative incidence rates at the start of the pandemic in 2020 (79, 81, 83). However, following the first lockdown, an increase in the administrative incidence of certain mental and behavioural disorders, surpassing pre-pandemic levels, was recorded (78, 83). Psychotherapists also reported a heightened demand for psychotherapy and pandemic-associated deteriorations in mental health (44, 88–91).

#### 4.2.3 Availability and Accessibility of Psychotherapeutic Care

Waiting times for treatment are often used as a marker of the availability of care. In the studies we reviewed, we found high variance in waiting times for an initial consultation and guideline-based therapy both between regions and between individual practices. This disparity is concerning in a healthcare system founded on the principle of solidarity, which aims to ensure equal access to care nationwide. Furthermore, chances of even receiving an appointment are unacceptably low, with waiting times reported to be excessively long and having doubled during the COVID-19 pandemic (41, 42, 44, 92). Psychotherapists reported receiving five to six requests for an initial appointment per week, of which only one-third resulted in an appointment (41, 42, 92). Half of the patients waited longer than four weeks for an initial consultation and more than six months for the start of guideline-based psychotherapy (41, 42). Given the time-sensitive development of children and adolescents, these difficulties in obtaining an appointment, followed by extensive waiting times, can lead to an exacerbation of symptoms, impaired psychosocial development, and damage to social participation. Additionally, they can result in reduced trust in the healthcare system and decreased motivation to seek professional help.

Research on individual barriers to accessing psychotherapeutic care for children and adolescents is still lacking. Nevertheless, we found indications that limited knowledge about mental health and mental healthcare services, fear of stigma, and negative experiences in the healthcare system are associated with a reduced likelihood of attempting to access psychotherapeutic care (56). Gender, socio-economic status, specific diagnosis, and severity of mental health problems and related impairment seem to moderate the motivation to access psychotherapeutic care (57, 58).

Taken together, we found a wide range of data on psychotherapeutic care for children and adolescents in Germany, allowing for some conclusions about the need and demand, service use and provision, and availability and access to care. It seems that the number of children and adolescents receiving psychotherapeutic treatment is far smaller than the number estimated to have an objective or subjective need for this treatment. While the demand for psychotherapeutic care appears to be rising in light of global crises, the capacities of psychotherapeutic care have not been adequately adjusted, leading to impaired access to care and long waiting times.

However, these conclusions should be interpreted with caution, as we had to compare results from different data sources which vary greatly in their content, methodology, and study purposes. The current database is very fragmented, and the data available on specific constructs is often very limited. Systematic, multimodal, comprehensive, and longitudinal assessments are necessary to reliably assess the state of the psychotherapeutic care system, which unfortunately do not exist.

### 4.3 Methods to Assess Outpatient Psychotherapeutic Care

In the following, we discuss the suitability of different data sources used to assess the outpatient psychotherapeutic care for children and adolescents in order to answer our third research question. The outcomes, main strengths, and limitations of each data source are summarised in Table 7.

**Table 7.**
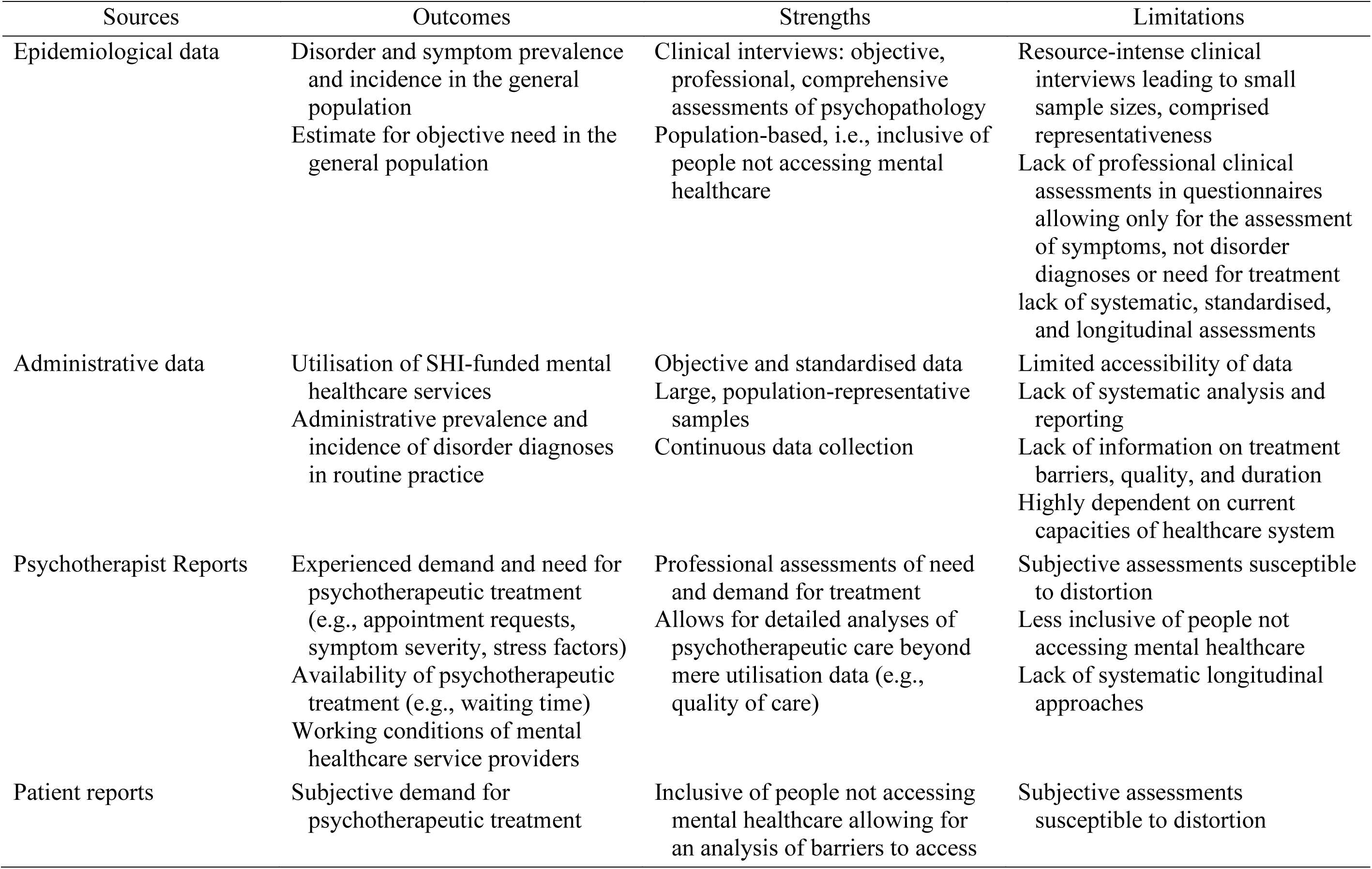
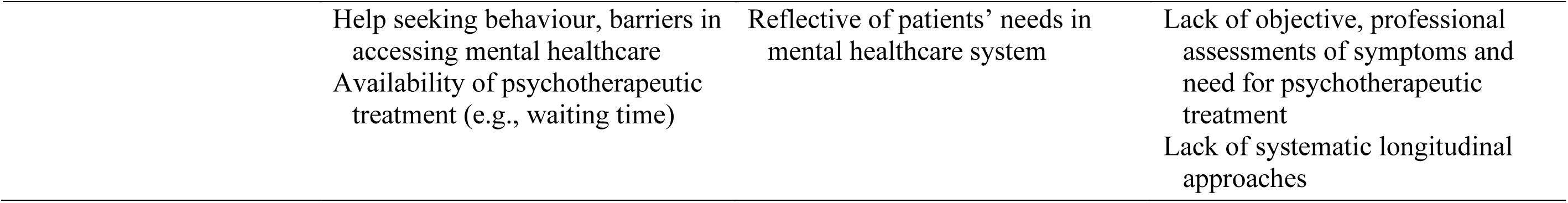
Summary of Possible Outcomes, Main Strengths and Limitations of Different Data Sources.

#### 4.3.1 Epidemiological Data

Epidemiological data can be used to determine the prevalence and distribution of mental and behavioural disorders and the need for treatment. Epidemiological studies typically use clinical interviews with children and adolescents to assess for mental or behavioural disorder diagnoses. Children, adolescents, or caregivers might also be asked if they have received a mental or behavioural disorder diagnosis in the past. Frequently, epidemiological studies also use questionnaires to assess general or disorder-specific psychopathological symptoms via self-report or other-report. These questionnaires can indicate whether the children, adolescents, or their caregivers observe symptoms that might be clinically relevant and require professional clarification.

A major advantage of using epidemiological data in assessing mental healthcare in comparison to data from routine practice is that it provides an objective estimate of the prevalence of specific mental or behavioural disorders in the general population. Therefore, it can be used to assess the objective need for professional mental healthcare based on treatment guidelines for respective disorders. Furthermore, if the epidemiological data is derived from clinical interviews conducted by professionals, it might serve as relatively objective, professional, and comprehensive assessments of psychopathology and need for treatment, being less prone to distortion than other data sources (e.g., patient reports).

However, epidemiological data alone is not sufficient to assess the state of mental healthcare provision as it most often does not include information on treatment rates but rather serves as a marker for the need and demand for treatment. Epidemiological data itself holds several limitations: Firstly, uniform definitions and standardized approaches for the assessment of many mental or behavioural disorders in childhood and adolescence are still missing (95, 98). Thus, there might be restrictions in the validity of assessed symptoms or diagnoses due to inconsistencies in case definition, assessment tools, and combination of different informants (95). While clinical interviews are the gold standard to diagnose mental and behavioural disorders, they are time- and resource-intensive and therefore seldom used in larger, representative samples in epidemiological studies. Large samples are, however, necessary to adequately assess less prevalent syndromes and access to mental healthcare as they are only relevant to a smaller subgroup of the sample (95). The more frequently used questionnaires alone are, however, not suited to determine clinical diagnoses requiring treatment, as a rather artificial cut-off score in a questionnaire does not equal the presence of a clinically relevant disorder (99). The results in a clinical questionnaire might also be distorted by the subjective biases of the children, adolescents, and caregivers due to factors such as stigmatization or mental health literacy (98). The assessment of mental health problems in younger children with clinical questionnaires is particularly challenging as younger children’s ability to reflect and report on symptoms is still limited, and especially internalising symptoms often go unnoticed by the caregivers. Furthermore, clinical question-naires often differ in researched symptoms, case definitions, indicators, operationalization, and reference population, leading to variance in data (98). Additionally, to assess changes in pathology due to developments in the healthcare system, longitudinal studies are necessary, which are, however, lacking.

In the study sample included in the current review, we found a lack of studies using clinical questionnaires and studies specifically assessing the need for psychotherapy, particularly in younger children. The latter would be particularly important as the prevalence of mental or behavioural disorder symptoms does not equal the need for psychotherapy. Furthermore, many of the included studies were associated with the same project (KiGGS-/BELLA-/COPSY-study). On the one hand, this leads to better comparability between the studies due to similarities in methodology and allows for longitudinal assessments. On the other hand, this accumulation of studies from the same project constricts the data to the research focus of the specific project (i.e., health-related quality of life, general measures of mental health problems). In the other studies, we found a large variety in assessed symptoms, limiting the options for comprehensive conclusions.

#### 4.3.2 Administrative Data

Administrative data comprises standardised billing information from healthcare service providers on the utilisation of healthcare services. The data can be provided by health, pension, or accident insurance agencies or by hospital statistics. In this review, we focus on data derived from SHI agencies.

A strength of this data source is its high ecological validity, as it is directly derived from routine practice. Additionally, administrative data provides an objective and standardized description of healthcare service provision. In Germany, around 90% of the population has SHI, hence the data from SHI agencies provides large and approximately representative samples of the population. Therefore, this data is less susceptible to distortion due to selection biases compared to epidemiological studies, patient reports, or therapist reports. Due to the large amount of available data, both in terms of sample size and number of assessed variables, a wide range of population-based analyses are possible (47, 81–83). Information which can be drawn from SHI data includes diagnosis and service data from SHI-accredited medical, therapeutic, pharmaceutical, hospital, and rehabilitation services (48, 81–83). This data provides information on prevalence rates of coded diagnoses, frequency of utilization of certain healthcare services, and waiting times between services. It can also include information on caregivers’ incapacity to work and (child) sickness benefit payments due to certain mental or physical disorders (47, 81–83). Moreover, administrative data can be collected continuously, allowing for the longitudinal analysis of trends in service provision and coded diagnoses.

However, these analyses are often limited because administrative data is typically published in reports by SHI agencies rather than empirical studies. Thus, the data presentation can be distorted by the interests of the SHI agencies. The agencies often publish focus analyses on specific aspects of children’s and adolescents’ mental health (e.g., specific diagnoses or billing codes) and do not provide comprehensive reports (81–83). Most often complete data and conducted analyses cannot be openly accessed. Further, there is a lack of standardised operationalisation and reporting as well as systematic statistical evaluations. This leads to variance in the definition of core variables between studies. For example, there are differences in the criteria for defining “cases” of mental or behavioural disorders (e.g., in how many quarters of a year a respective diagnosis has to be documented, which medical service has to be billed). Additionally, reports differ in which services (i.e., billing codes) are counted as “psychotherapeutic care” or “mental healthcare.” Some reports do not even specify how cases or mental healthcare are conceptualized. These definitions, however, significantly influence the results and limit comparability between reports (52).

Regarding diagnosis prevalence rates, it must be noted that the quality of diagnostic evaluation in routine practice is often poor, as it highly depends on the assessment of the respective practitioner and is not linked to a specific diagnostic procedure (100). Therefore, the validity of the coded mental or behavioural disorder diagnoses is limited (86). Furthermore, the prevalence and incidence rates of coded diagnoses are highly dependent on the current capacities of the mental healthcare system, which is restricted by demand planning. The actual demand for diagnostic evaluation and psychotherapeutic care, as well as changes in demand, cannot be depicted in administrative data since the capacities for SHI-funded outpatient psychotherapeutic care are already fully utilized. It can be assumed that not all children and adolescents in need of diagnostic evaluation or care are able to access the system. Administrative data is not suited to assess barriers to treatment access and does not include information on children and adolescents not entering the healthcare system. Regarding psychotherapeutic care itself, there is a lack of information on the type, duration, quality, and effectiveness of treatment in administrative data.

Consequently, it is important to keep in mind that administrative data is not a valid indicator for epidemiological prevalence and incidence rates, the demand for psychotherapeutic treatment, or adequate psychotherapeutic care. It can merely depict the services and diagnoses billed in the current SHI-funded healthcare system.

In this review, we found both reports published by respective SHI agencies and more standardized empirical studies published by the German Central Institute for SHI Physicians. These publications reported on a wide range of outcomes, including prevalence and incidence rates of documented mental or behavioural disorders, frequency and modalities of treatment, and changes in diagnosis and treatment prevalence during the COVID-19 pandemic. Six reports were published by the same SHI agency in a series (“DAK Gesundheitsreport”). However, these reports vary greatly in their focus (specific diagnoses, prevalence rates, incidence rates), definitions of treatment, and the time spans they cover (e.g., data from one year vs. multiple years, data from one year or up to five years before the report). This makes it difficult to compare these publications, even though they belong to the same series. Additionally, the included publications do not differentiate between mental and behavioural disorders requiring different types of treatment. Regarding psychotherapy, it must be considered that diagnoses from the diagnostic groups ICD-10 F7 and F8 are mostly not an indication for psychotherapy. Thus, the overall prevalence of any ICD-10 F-diagnosis in the data cannot be used as an estimate for the need for psychotherapeutic care.

#### 4.3.3 Psychotherapist Reports

Reports on the current state of outpatient psychotherapeutic care from the perspective of providers in private practice (including child and adolescent psychotherapists, psychological therapists, and medical psychotherapists) are often based on online surveys conducted by professional associations or psychotherapist chambers.

This data can provide professional evaluations of current stress levels and stress factors in children and adolescents, thereby contributing to assessments of the need for treatment. Psychotherapists can further report on the demand for diagnostic evaluations or psychotherapeutic treatment they experience, including requests for appointments that cannot be fulfilled. Therefore, these studies can provide more information on patients seeking help but unable to access the healthcare system than epidemiological or administrative data. Additionally, they can provide estimates of waiting times for initial consultations and the start of guideline-based psychotherapy and information on the treatment formats psychotherapists offer and the disorders they are treating. Furthermore, psychotherapist reports can offer unique insights into the experiences of psychotherapists as healthcare providers, such as their working conditions, stress levels, and stress factors. This information is crucial for a comprehensive evaluation of the psychotherapeutic healthcare system, as adequate treatment is only possible when service providers have decent working conditions.

Nevertheless, these studies are susceptible to distortion due to the subjective assessments of the psychotherapists, which can be influenced by individual perceptions, assumptions, attitudes, and stress, as well as certain biases or errors in reporting (e.g., hindsight or memory bias). This might limit the validity and reliability of the derived data. Moreover, in an exhausted healthcare system, there might be selection biases in the samples of participants, potentially reducing the representativeness of the data.

In the articles we reviewed, we found that multiple surveys do not include or do not report separately on psychotherapists who work with children and adolescents and therefore had to be excluded, limiting the available data.

#### 4.3.4 Patient Reports

Patient reports in our review included data derived from surveys among children, adolescents, or their caregivers on the children’s and adolescents’ demand for and access to mental healthcare.

This data can include self- or caregiver-perceived mental health problems, the utilization of professional help, the demand or motivation to seek such help, and barriers experienced in accessing the healthcare system. While epidemiological data and psychotherapist reports can serve as measures of the objective need for psychotherapeutic care, patient reports can serve as a measure of subjective demand for this treatment. An advantage of this data source over others is that it provides insights into children and adolescents who may need psychotherapeutic treatment but do not do not succeed in accessing it.

However, it should be considered that these studies lack professional assessments of symptoms and the need for treatment, which can limit the validity of the data. Furthermore, assessing symptoms and the need for psychotherapeutic care is especially difficult in younger children due to their still-limited ability to self-reflect and communicate. Therefore, studies often use caregiver reports, which can be distorted by the caregivers’ perceptions, attitudes, and stress. Moreover, the assessment of need or use of different kinds of mental healthcare (e.g., treatment by a psychiatrist, psychologist, or psychotherapist) is limited by the participants’ knowledge and ability to correctly differentiate between these different healthcare providers.

In our data we found that patient reports on perceived demand for mental healthcare often confounded with epidemiological studies. In the studies included in this category, “mental healthcare” was not further specified, which does not allow for statements on psychotherapeutic care in particular. Overall, we found a lack of studies using patient reports and a lack of data on quantity, quality, and access to mental healthcare.

In conclusion, it becomes evident that each data source provides unique insights into the psychotherapeutic care situation. However, relying on a single data source is insufficient, as each has limitations in the scope of variables it can assess and the quality of the data it provides. Therefore, a comprehensive assessment requires considering multiple data sources to capture indicators of both objective and subjective need and demand for psychotherapeutic treatment, as well as factors such as utilization, quantity, quality, accessibility, and availability of care.

### 4.4 Strengths and Limitations of the Current Review

To our knowledge, the current review is the only comprehensive study in recent years that examines the psychotherapeutic care situation for children and adolescents in Germany using a variety of data sources. However, due to regular changes in the healthcare system, demand planning, and current societal challenges potentially influencing the mental health of children and adolescents (e.g., the COVID-19 pandemic, the climate crisis, global conflicts), it is necessary to continuously collect and discuss current data on the provision of psychotherapeutic care. A strength of this review is that it provides not only an overview of recent data, but also a methodological discussion of the strengths and limitations of different data sources. Furthermore, we included empirical studies as well as administrative reports, thereby offering a combination of scientific research and practical care data, thus linking research and practice. Finally, our extensive search covered four databases and included grey literature, reducing the influence of publication bias on our results.

This work, however, has several limitations. Most importantly, we focused solely on the provision of outpatient psychotherapeutic care, which is by far not the only healthcare service for children and adolescents with mental or behavioural problems in Germany. Other professions involved in the mental healthcare of children and adolescents are paediatricians, child and adolescent psychiatrists, social workers, psychologists, teachers, educators, occupational therapists, and child and youth welfare professionals. Therefore, children and adolescents who do not access outpatient psychotherapeutic treatment might still receive treatment from other professions or in different settings. For example, Germany has the highest number of inpatient beds for children and adolescents in Europe, indicating that many children and adolescents with mental or behavioural disorders could receive mental healthcare in the inpatient sector (34). Moreover, our data does not allow for conclusions on the quality or duration of psychotherapeutic treatment but rather on the quantitative demand, service use, and access.

Methodologically, while including peer-reviewed empirical articles, administrative reports, and grey literature broadens our data scope, it also introduces potential limitations in the quality of included publications, which we did not assess or account for. We did not statistically evaluate the size of the publication and reporting bias, so we cannot estimate their effects on our data. Due to the scoping approach of our review, we found significant heterogeneity in the data regarding data sources, methodology, and reported outcomes. This heterogeneity prevents a quantitative summary of the data. Thus, our results are based on a qualitative synthesis of the research findings. Lastly, the lack of epidemiological studies on the prevalence rates of mental and behavioural disorders in children and adolescents and on patient reports limits our ability to draw conclusions on this data, leaving some research questions insufficiently answered.

### 4.5 Practical Implications

Our results suggest that approximately 13–17% of German children and adolescents are be in need of psychotherapeutic care, while only 5–7% receive broader mental healthcare and only 0.2–2% receive guideline-based psychotherapy. Given the serious individual and societal consequences that untreated mental and behavioural disorders in childhood and adolescence can have, this discrepancy is alarming. One reason children and adolescents may not access outpatient psychotherapeutic care could be a lack of knowledge about mental health, mental healthcare services, and how to access them, as well as prejudice and stigma surrounding psychotherapeutic care among children, adolescents, and their families. To address these barriers, interventions are needed to provide education on mental health, increase awareness of mental healthcare services, and combat stigma associated with mental health and psychotherapy, particularly in hard-to-reach populations.

Another major reason is the limited availability of SHI-funded outpatient psychotherapy. Compared to other high-resource countries, such as Italy, the Netherlands, or the USA, it seems that psychotherapeutic care in Germany is behind in terms of the ratio of mental health professionals to the population and the number of children and adolescents accessing mental healthcare (3, 38).

It is clear that the current capacity of psychotherapeutic care is not sufficient to meet the need for treatment. This is reflected in the large proportions of unmet requests and long waiting times for treatment, which are an immense burden on families with mentally ill children and adolescents who are willing to seek professional help. This problem only seems to be becoming more drastic in the future, as the demand is predicted to rise even further due to de-stigmatisation and improvements in early diagnostics of mental and behavioural disorders, as well as an increase in stress factors for children and adolescents (e.g., the advancing climate crisis, escalating global conflicts, increasing academic pressure, and the growing shortage of teachers in Germany) (101). The capacity of SHI-funded outpatient health services is determined by the system of “Demand Planning” carried out by the Federal Joint Committee, the National Association of Statutory Health Insurance Funds, and the National Association of Statutory Health Insurance Physicians at the federal level. It determines how many practitioners of a certain medical profession (here psychotherapy) are allowed to provide health services at the expense of SHI funds in a certain area. These numbers are, however, not based on epidemiological calculations of actual demand for services but merely on the continuation of a certain historical ratio between practitioners and the population (49–51). Moreover, there is currently no independent demand planning for child and adolescent psychotherapy. The numbers of SHI-accredited psychotherapists for children, adolescents, and adults are planned together, with a rather artificial quota for child and adolescent psychotherapy. Hence, the current system has been heavily criticised, and multiple expert reports and statements from psychotherapist organisations have called for major reforms over the last few years (49, 50, 102–111).

The results of the current review strongly support the need for a reform in demand planning, as data clearly indicates that the current number of SHI-accredited psychotherapists is not sufficient to adequately and timely provide the necessary amount of psychotherapeutic care for children and adolescents in Germany. An urgent expansion of SHI-funded psychotherapeutic care capacity is needed to protect the long-term mental health and social participation of children and adolescents and to prevent significant societal costs associated with untreated or late-treated mental and behavioural disorders, such as increased treatment expenses, sick leave costs, and early retirement.

### 4.6 Implications for Future Research

We have demonstrated that a comparative analysis of different data sources is necessary to comprehensively assess outpatient psychotherapeutic care for children and adolescents. However, our review revealed significant research gaps, particularly concerning epidemiological data on mental and behavioural disorders in German children and adolescents, and patient reports on access to psychotherapeutic care. Therefore, further research focusing on these areas and integrating various data sources is needed.

To adequately assess changes in the mental health of children and adolescents over time, longitudinal study designs are essential. The KiGGS-/BELLA-/COPSY-study is one example of such a longitudinal epidemiological research project. Continuous funding and support for these projects are crucial to obtaining reliable data on child and adolescent mental health in the context of evolving challenges and social crises. Similarly, systematic, comprehensive, and longitudinal study designs are necessary to assess the psychotherapeutic care situation from the perspectives of psychotherapists and patients. Regarding administrative data, our review highlighted that systematic and standardized approaches to analysing and reporting this data are still lacking. Such standardization is essential for effective research and service planning, as it allows for meaningful comparisons and analysis of changes over time, ensuring that this data source can be fully utilized.

International studies on psychotherapeutic care provision yield similar conclusions. A comparable review from Austria, which assessed epidemiological studies and administrative data to estimate prevalence rates and care provision, found a lack of epidemiological studies in German-speaking countries and great variance in data (112). The authors concluded that systematic data is necessary to comprehensively assess the demand for and provision of psychotherapy. Furthermore, an international survey assessing the provision of child and adolescent mental health services in 28 European countries revealed that a major challenge shared among countries is the lack of systematic and standardized assessments of service provision and quality (34). This lack of systematic structures to comprehensively assess the psychotherapeutic care situation appears to be an international issue, not just a national one.

Lastly, as discussed in the limitations of this review, mental healthcare is also provided by other professions and in the inpatient sector. Exploring the provision of care in these areas could offer valuable insights and lead to more comprehensive conclusions about the overall healthcare system.

## 5 Conclusion

This scoping review of 41 publications on the state and assessment of SHI-funded outpatient psychotherapeutic care for children and adolescents in Germany indicates that approximately one in four to five children has a mental or behavioural disorder, and one in six to seven children requires psychotherapeutic treatment. This need is largely unmet, as the majority of treatment requests are not fulfilled, waiting times for therapy are excessively long, and only up to 10% of children and adolescents with a mental or behavioural disorder receive guideline-based psychotherapy. These findings underscore the insufficiency of current treatment capacities to meet the increasing mental healthcare needs of children and adolescents and highlight the urgent need for expansion to prevent long-term harm to current and future generations. Our findings emphasize the necessity for a systematic, multimodal, and longitudinal analysis of the care system. Such an analysis should integrate epidemiological, administrative, psychotherapist, and patient data to empirically assess the need and demand for treatment, service use and provision, and the availability and accessibility of care. Future research must provide these comprehensive and systematic assessments to scientifically guide and evaluate changes in the mental healthcare system.

## 6 Conflict of Interest

The authors declare that the research was conducted in the absence of any commercial or financial relationships that could be construed as a potential conflict of interest.

## 7 Author Contributions

Idea: JS; Conceptualisation: KRW, JS; Methodology: KRW; Literature search and data analysis: KRW; Writing – original draft: KRW; Writing – review & editing: KRW, JS; Supervision: JS.

## 8 Funding

This review was funded by the Robert Bosch Stiftung GmbH (grant number: 01000971-001). The funders had no role in study design, data collection and analysis, decision to publish, or the preparation of the manuscript. The publication was supported by the Open Access Publishing Fund of Leipzig University.

## Data Availability

All data produced in the present work are contained in the manuscript.

## Acknowledgements

The authors would like to acknowledge that the language editing for this manuscript was supported by ChatGPT (GPT 3.5), an artificial intelligence language model by OpenAI, to enhance the readability and clarity of the text without altering or generating the scientific content.

## Notes

### Competing Interest Statement

The authors have declared no competing interest.

